# Local Influenza Forecasts Outperform State-Level Forecasts in the United States

**DOI:** 10.64898/2026.06.04.26354836

**Authors:** Dongah Kim, Remy Pasco, Kaitlyn Johnson, Spencer J. Fox, Nicholas G. Reich, Lauren Ancel Meyers

## Abstract

Accurate outbreak forecasts are critical for timely and effective public health response. In the United States, however, most forecasts are produced at the state level, which can mask substantial sub-state heterogeneity and limit their utility for local planning. We generated and evaluated forecasts of the percentage of Emergency Department visits attributable to influenza across 173 large metropolitan Health Service Areas (HSAs) using a gradient boosting quantile regression (GBQR) model, and compared their accuracy to forecasts derived from state-level data alone. At a one-week, two-week and three-week horizon, local forecasts outperformed state-based forecasts in 98.8%, 90.8%, and 78.6% of HSAs, respectively, achieving mean weighted interval scores that were on average a 39.2% lower (95% range: 5.9% to 76.7%), 19.6% lower (−6.3% to 59.5%), and 11.4% lower (−11.7% to 44.9%), respectively. The performance advantage of local forecasting was strongest in HSAs representing a smaller share of their state’s population and increased with the proportion of the HSA population living in urban areas and the number of metropolitan areas within a state. These results, based on an analysis of HSAs with populations greater than 250,000, demonstrate that fine-scale modeling can substantially improve forecast accuracy and highlight the potential value of local forecasts for outbreak preparedness and response.

**Significance Statement:** During severe influenza seasons and pandemics such as COVID-19, anticipating the timing and severity of local peaks is critical for managing healthcare surges and communicating risk—yet these dynamics can vary substantially across local areas within the same state. However, most forecasting efforts in the United States provide only state-or national-level projections. This study shows that local influenza forecasts are both feasible and consistently more accurate than state-level projections, particularly in metropolitan areas that make up a small share of large states with multiple metropolitan areas.

## Introduction

Infectious disease forecasting has become a vital tool for public health planning, with its importance growing steadily over the past two decades (1–5). The COVID-19 pandemic further underscored its value, as forecasting models were widely used to guide healthcare surge management and critical interventions such as social distancing, mask mandates, and vaccination strategies. Starting in 2020, dozens of modeling groups submitted weekly forecasts of state and national-level cases, hospitalizations, and deaths to the COVID-19 Forecast Hub, providing situational awareness and decision support to policymakers (6, 7). Beyond COVID-19, forecasting has been applied to a wide range of pathogens—including influenza, RSV, dengue, cholera, Ebola, and measles—to anticipate epidemic peaks, allocate limited healthcare resources, and plan timely public health messaging (8–14).

Over the past decade, the infectious disease modeling community has established a robust collaborative infrastructure to support real-time forecasting (15). Initiatives such as the CDC’s FluSight Challenge (16) and the COVID-19 Forecast Hub (17) have demonstrated the utility of ensemble modeling (9, 18), transparent data sharing (19), and systematic evaluation frameworks (7, 20). These efforts have accelerated methodological innovation, integrating statistical, mechanistic, and machine-learning approaches that leverage new data streams such as mobility, vaccination coverage, and climate indicators. As a result, epidemic forecasting is increasingly viewed as an essential component of public health preparedness and response.

Despite these advances, a critical gap remains in translating insights from national and state-level forecasts to the local scale. In the U.S., public health responses—such as vaccine distribution, school closures, and outreach campaigns—are often implemented at sub-state levels, including cities, counties, or health regions (2). However, state-level forecasts often mask important geographic heterogeneity in disease dynamics (21, 22). This limitation is especially pronounced in large states such as Texas, where metropolitan areas like Houston and Dallas experience distinct epidemic trajectories, driven by differences in demographics, mobility patterns, and healthcare infrastructure. Beyond heterogeneity, this discrepancy creates an unequal forecasting landscape, in which residents of and decision-makers in physically small states benefit from geographically precise forecasts, while those in larger states must rely on highly aggregated forecasts.

Recent advances in data infrastructure and computational modeling now make local-level forecasting both feasible and timely. The CDC’s National Syndromic Surveillance Program (NSSP) provides near-real-time emergency department (ED) visit data across the U.S., capturing patterns of viral respiratory illness at high spatial and temporal resolution (8). These data offer a unique opportunity to systematically evaluate the value of local-scale forecasts. Modern statistical and mathematical modeling techniques enable scalable and interpretable forecasting across hundreds of local regions, while maintaining statistical rigor and uncertainty quantification.

A growing body of work has demonstrated the feasibility and operational value of infectious disease forecasting at the sub-region level (23). For seasonal influenza, Shaman and colleagues conducted real-time, ensemble-based forecasts for 108 U.S. cities, showing that reliable predictions of key outbreak features, such as peak timing, are achievable at city resolution (14, 24). Subsequent studies extended these efforts by evaluating and combining multiple forecast systems across many U.S. cities, including superensemble approaches, further supporting the routine generation and comparison of local-scale forecasts (4, 6, 7). More recently, hierarchical models have been used to generate short-term influenza hospitalization forecasts across English subregions, demonstrating operational utility for public health planning (25). At the same time, other work suggests intrinsic limits to forecasting at finer spatial scales. White & León (26) show that forecast error increased in locations with smaller populations, implying greater stochastic noise and weaker signal in less populous jurisdictions. This raises an important question: while local forecasts are clearly valuable, can they be produced with consistent reliability across diverse jurisdictions and surveillance conditions, or are state-level forecasts, when applied to local areas, more reliable?

Evidence from multi-model FluSight evaluations provides a more nuanced picture. Although performance varies substantially across U.S. states, published analyses do not show a simple monotonic relationship between forecast accuracy and population size (1, 27). Together, these findings suggest that while smaller jurisdictions may pose structural challenges to models seeking to make highly accurate forecasts, realized forecast performance depends on additional epidemiological, surveillance, and modeling factors beyond population size alone.

This study evaluates the value of local-level infectious disease forecasting compared to existing state-level forecasts. Specifically, we assess whether influenza forecasts at the level of Health Service Areas (HSAs)—government-defined healthcare catchments (28)—are more accurate when generated using local HSA-level data or state-level data alone. Using viral respiratory illness data from the CDC’s National Syndromic Surveillance Program (NSSP) (29), we compare short-term forecasts of the percent of ED visits due to influenza generated at the local (HSA) and state scales, with both forecasts evaluated at the HSA level, across several standard accuracy metrics: prediction interval coverage, mean absolute error (MAE), and mean weighted interval score (MWIS) (20), and identify robust predictors of where local forecasting adds the greatest value.

## Results

Using data from the CDC’s National Syndromic Surveillance Program (NSSP), we generated forecasts of the percentage of emergency department (ED) visits attributable to influenza for multi-county Health Service Areas (HSAs) across the continental United States with publicly available data from October 2022 through July 2025. After excluding HSAs with populations below 250,000, Washington, DC, and HSAs with incomplete data, the final analysis included 173 HSAs across 31 states.

We compared the percentage of influenza-related ED visits at the state and HSA levels. Large discrepancies between these time series suggest that local and state forecasts may diverge. To illustrate, we highlight HSAs in Georgia, New York, North Carolina, and Texas in which local ED visit trends differ sharply in timing and intensity from their state aggregates (Figure 1A). Supplementary Figure A.S1 gives corresponding plots for the 20 HSAs with the largest and smallest local-state discrepancies.

**Figure 1.**
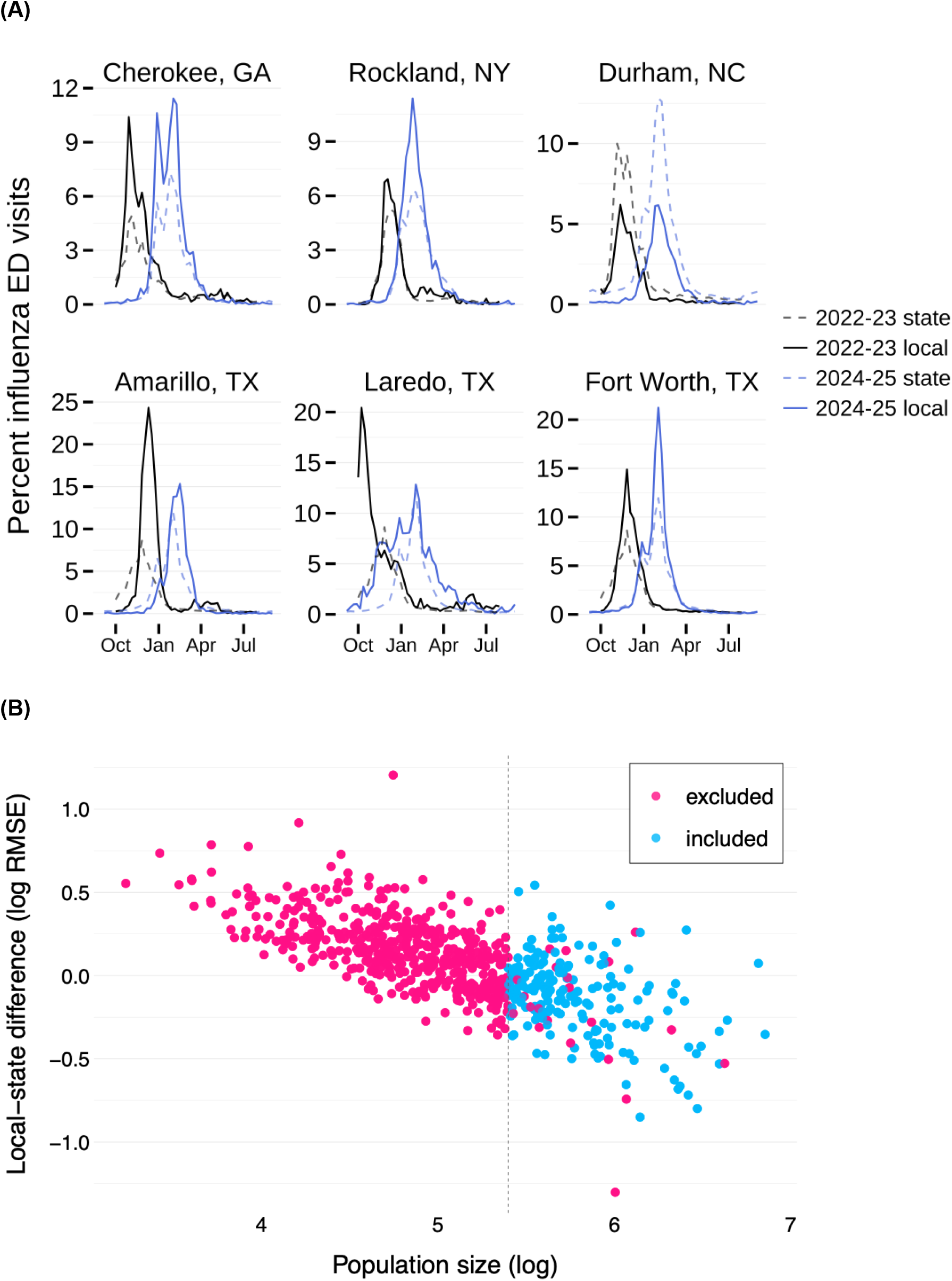
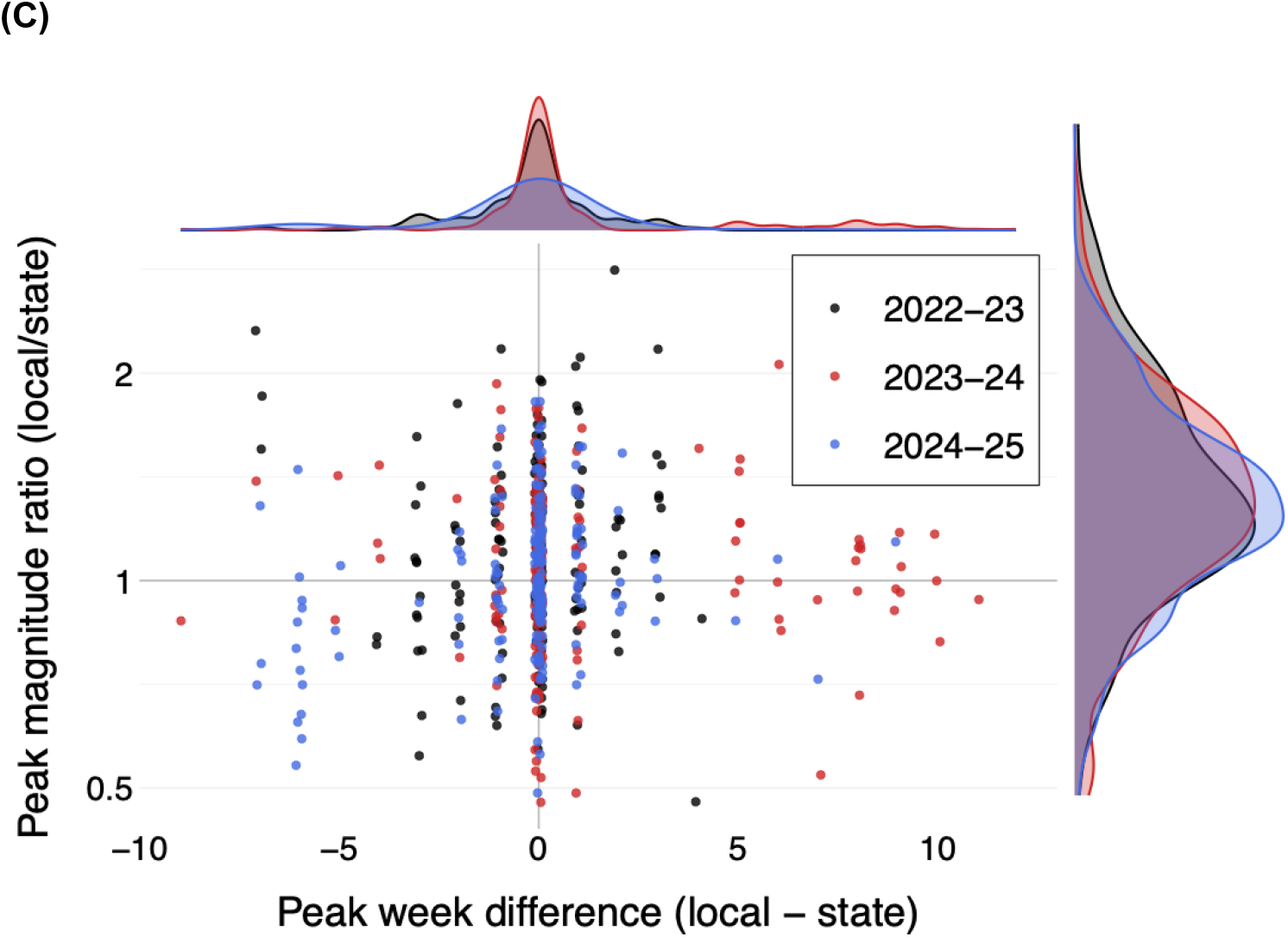
Comparison of local- and state-level trends in the percentage of emergency department (ED) visits attributable to influenza during the 2022–2023, 2023–2024, and 2024–2025 influenza seasons. (A) Six HSAs selected as illustrative extreme examples showing large discrepancies between local- and state-level influenza trends. The labels above each graph correspond to the largest city in the multi-county HSA, except for Rockland, which represents a single-county HSA. Lighter dashed lines indicate state-level trends; darker solid lines indicate the corresponding HSA-level trends. (B) Relationship between the local-state difference (root mean squared error [RMSE]) and HSA population size. The set of HSAs included is limited by the availability of publicly reported NSSP influenza trend data. Forecast evaluation was further restricted to HSAs with populations over 250,000 and to those with sufficiently stable time series, excluding regions with extremely noisy data or those outside the continental United States (shown in blue). (C) Differences in the timing and magnitude of local versus state influenza peaks across seasons. The x-axis shows the timing offset (local peak week minus state peak week), and the y-axis shows the ratio of peak magnitudes (local peak percentage divided by state peak percentage). Because the y-axis is displayed on a logarithmic scale, equal distance above and below 1 represent symmetric relative differences (e.g., ratios of 2 and 0.5 appear equally distant from 1). Each point represents an HSA- season pair, colored by influenza season. For example, the leftmost point (red) from the 2023–2024 season corresponds to the forecast for HSA 546 (the region around Des Moines, IA); the local influenza trend peaked 9 weeks earlier than the state-level trend, with a peak 13% lower than the statewide peak.

Using root mean square error (RMSE) to quantify similarity between local and state trends, we found an inverse relationship between HSA population size and local–state agreement. Smaller HSAs exhibited larger deviations from state-level trends (Figure 1B), reflecting both true local heterogeneity and increased week-to-week stochastic variability associated with smaller populations. To reduce stochastic noise, we restricted subsequent analyses to HSAs with populations exceeding 250,000 (Supplementary Material B). RMSE values for all 173 HSAs are provided in Supplementary Table A.S5, with geographic patterns shown in Supplementary Figure A.S8.

Although RMSE captures overall divergence between local and state trajectories, the largest discrepancies often occurred near epidemic peaks—periods when accurate forecasts are most valuable for healthcare planning and public communication. On average, local peak timing and magnitude resembled statewide trends. However, substantial variation existed across locations and seasons, with some local peaks occurring more than five weeks earlier or later and reaching up to 2.8-fold higher or lower intensity than corresponding state peaks (Figure 1C).

We applied the Gradient Boosting Quantile Regression (GBQR) model (30) to generate retrospective forecasts for the 2022-2023, 2023-2024, and 2024-2025 influenza seasons. For each target season, models were trained on data from the other two seasons and on observations available up to the forecast date within the target season. GBQR, which performed strongly in recent CDC FluSight evaluations (16), generates full predictive distributions across quantiles, enabling evaluation using metrics such as Weighted Interval Score (WIS) and coverage rate (see Supplementary Material E). For comparison, we applied the same forecasting framework at both the state and HSA levels to two benchmark models: a quantile baseline producing flat forecasts and an automated ARIMA model. In general, the GBQR model outperformed both the ARIMA and baseline models on point and probabilistic accuracy metrics (MAE and MWIS), although its prediction intervals were slightly overconfident relative to ARIMA (Supplementary Table A.S1). Detailed model specifications are provided in Supplementary Material D.

We evaluated forecasting performance for 173 Health Service Areas (HSAs) across 1–4 week forecast horizons. As an illustrative example, Figure 2 shows forecasts for six focal HSAs selected because they exhibit large discrepancies between state-level and local epidemic dynamics during the 2023–2024 influenza season. Across these HSAs, the HSA-level 95% prediction intervals capture more of the observed ED visit trends than the corresponding state-level forecasts, particularly during epidemic peaks. In Cherokee, GA, Rockland, NY, and several Texas HSAs, state-level forecasts misestimate both the timing and magnitude of local peaks. Results for additional seasons and forecast horizons are provided in Supplementary Figure A.S2-A.S4. The main text highlights six HSAs with large HSA-state discrepancies in the observed data, while Supplementary Figure A.S5 shows corresponding results for six HSAs with small discrepancies.

**Figure 2.**
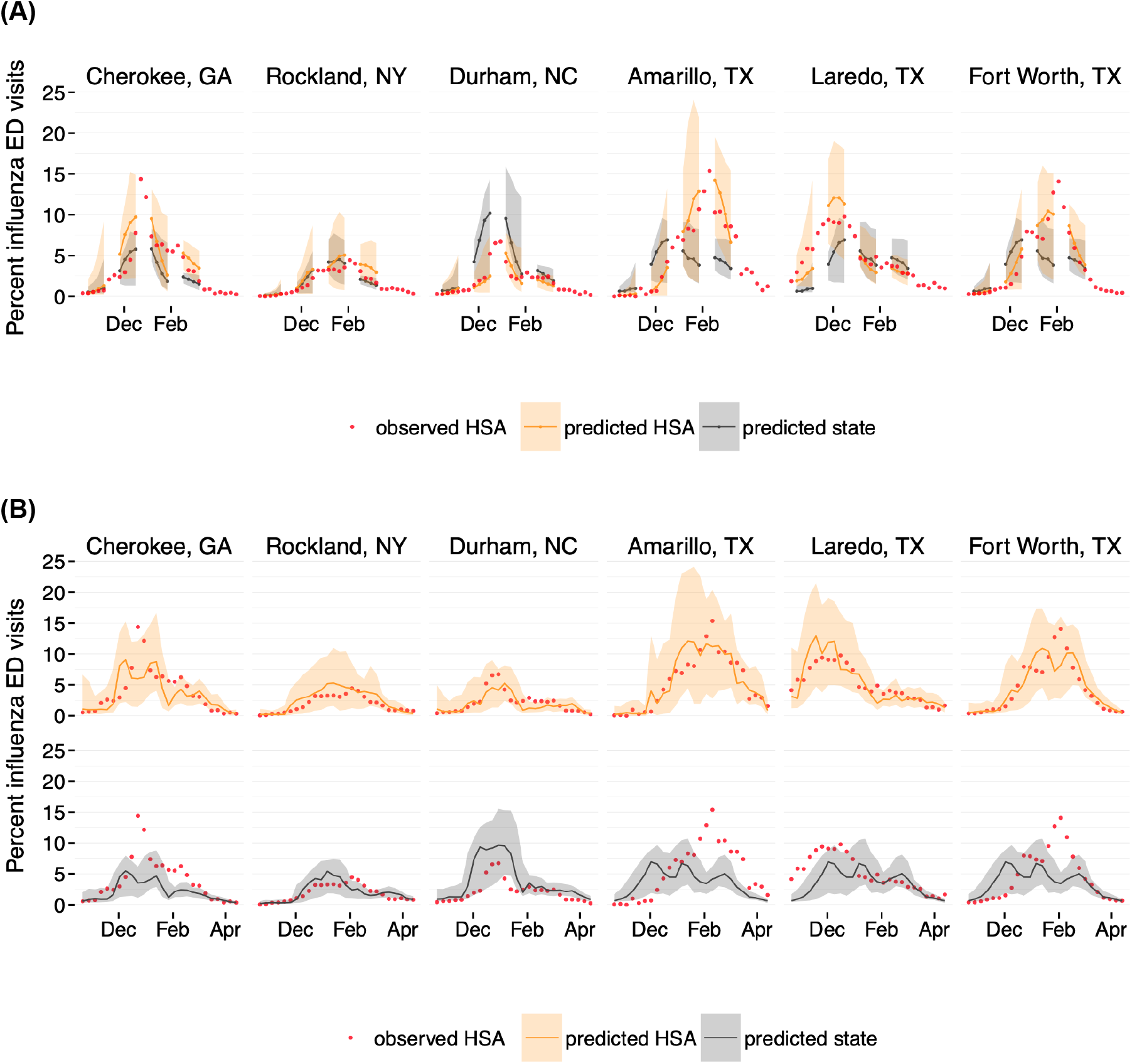
Forecasts of the percentage of emergency department (ED) visits attributable to influenza for six metropolitan Health Service Areas (HSAs) during the 2023–2024 influenza season. Red points indicate observed HSA-level ED visit percentages. Solid lines represent forecast medians, and shaded regions indicate 95% prediction intervals. Orange curves correspond to forecasts generated using HSA-level data, and gray curves correspond to forecasts generated using state-level data. (A) Four four-week-ahead forecasts initialized on four selected dates (October 14, 2023; November 25, 2023; January 6, 2024; February 17, 2024; and March 30, 2024) for each HSA. Within each panel, the 1-, 2-, 3-, and 4-week forecast trajectories from all four initialization dates are shown, with HSA-level forecasts in orange and state-level forecasts in gray, superimposed on the observed data. (B) Weekly three-week-ahead forecasts concatenated into a continuous time series. The top row shows forecasts generated using HSA-level data (orange), and the bottom row shows forecasts generated using state-level data (gray), each compared to observed HSA-level ED visits.

On average, HSA-level forecasts outperformed state-level forecasts across all forecast horizons. Local forecasts achieved lower MWIS values than state-level forecasts in 98.8%, 90.8%, 78.6%, and 69.4% of HSAs at the one-, two-, three-, and four-week horizons, respectively, with average MWIS reductions of 39.2%, 19.6%, 11.4%, and 6.5%. Although forecast gains diminished at longer horizons, average MWIS differences remained negative across all horizons considered. The distribution of HSA-specific differences in MWIS across forecast horizons is shown in Supplementary Figure A.S6. Consistent with this pattern, average performance metrics also favored local forecasts (Table 1). At the one-week horizon, HSA-level forecasts achieved a mean coverage rate of 0.91, compared with 0.59 for state forecasts, along with lower error. Nominal prediction interval coverage should approach 0.95 if forecast uncertainty is well calibrated; neither model reached this target, but local forecasts came substantially closer. Even at four weeks ahead, local coverage remained higher (0.72 vs. 0.58) and errors lower (MAE 1.39 vs. 1.44; MWIS 0.85 vs. 0.93). Similar patterns were observed when using a quantile baseline and an automated ARIMA model to generate state- and HSA-level forecasts (Supplementary Table A.S1). Season-specific comparisons across all models are provided in Supplementary Table A.S2-A.S4.

**Table 1.** Average forecast accuracy of HSA-level and state-level models across the 2022–2023, 2023–2024, and 2024–2025 influenza seasons. Performance is summarized for one-, two-, three-, and four-week-ahead forecast horizons. Metrics include 95% prediction interval coverage rate, mean absolute error (MAE), and mean weighted interval score (MWIS), each averaged across all locations and forecast weeks. Coverage rates closer to the nominal 95% level indicate better-calibrated uncertainty, while lower MAE and MWIS values indicate greater forecast accuracy. Relative differences for both MAE and MWIS were computed at the location level as (Metric_HSA_ - Metric_state_)/Metric_state_ and summarized using the mean and 95% range. “% HSAs exceed state” denotes the proportion of HSAs for which HSA-level forecasts outperformed state-level forecasts. Bold values indicate superior performance of HSA-level forecasts. Values in parentheses denote standard deviations across locations and forecast weeks for coverage, MAE, and MWIS, and 95% ranges for relative differences.

| Metric | Model | Forecast horizon |  |  |  |
| --- | --- | --- | --- | --- | --- |
|  |  | 1 week | 2 weeks | 3 weeks | 4 weeks |
| 95% PI Coverage Rate | HSA | **0.905 (0.293)** | **0.833 (0.373)** | **0.781 (0.413)** | **0.720 (0.449)** |
|  | State | 0.590 (0.492) | 0.614 (0.487) | 0.608 (0.488) | 0.580 (0.494) |
| MAE | HSA | **0.599 (0.851)** | **0.981 (1.356)** | **1.254 (1.622)** | **1.388 (1.744)** |
|  | State | 0.908 (1.284) | 1.147 (1.556) | 1.349 (1.726) | 1.439 (1.774) |
|  | Relative Difference | -0.290<br>(-0.668, 0.022) | -0.118<br>(-0.465, 0.125) | -0.057<br>(-0.347, 0.145) | -0.028<br>(-0.299, 0.179) |
|  | % HSAs exceed state | 96.0 | 76.3 | 61.3 | 53.2 |
| MWIS | HSA | **0.300 (0.443)** | **0.525 (0.817)** | **0.705 (1.079)** | **0.852 (1.281)** |
|  | State | 0.567 (0.959) | 0.692 (1.104) | 0.821 (1.255) | 0.929 (1.384) |
|  | Relative Difference | -0.392<br>(-0.767, -0.059) | -0.196<br>(-0.595, 0.063) | -0.114<br>(-0.449, 0.117) | -0.065<br>(-0.348, 0.207) |
|  | % HSAs exceed state | 98.8 | 90.8 | 78.6 | 69.4 |

Local forecasting requires additional data infrastructure and modeling effort, and its value may vary across regions. To identify where investment in local forecasting is most likely to yield meaningful gains over state-level models, we examined how improvements in forecast accuracy depend on population structure at both the HSA and state scales. We modeled average differences in weighted interval score for each HSA and *i* horizon *h* (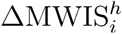; see Eq. (1)) between HSA- and state-level forecasts across all 173 HSAs as the outcome, with the HSA–state population ratio, the proportion of the HSA population living in urban areas, and the number of metropolitan statistical areas (MSAs) in the state as predictors, including an interaction term between population ratio and urbanization, using a generalized linear model (GLM). Here, the number of MSAs per state serves as a proxy for substate fragmentation in healthcare catchments and, by extension, potential heterogeneity in epidemic timing across regions. Partial residual plots from the fitted GLM show that among the larger HSAs for which we evaluated forecasts, those that represent a smaller fraction of their state’s population tend to experience larger improvements from local forecasting. Crucially, this effect is most pronounced in highly urbanized HSAs (Figure 3A), where the association between the HSA-state population ratio and forecast improvement is stronger across urbanization tertiles. Additionally, states subdivided into a larger number of MSAs also exhibit greater gains overall (Figure 3B). Estimated coefficients for all models are provided in Supplementary Table C.S1. While some more flexible modeling approaches showed modest improvements in predictive performance for certain forecast horizons, the overall results were broadly consistent across models. We report the results of the GLM here to prioritize interpretability and to transparently characterize how population structure relates to the added value of local forecasting. Results from more flexible models are provided in the Supplementary Material C.

**Figure 3.**
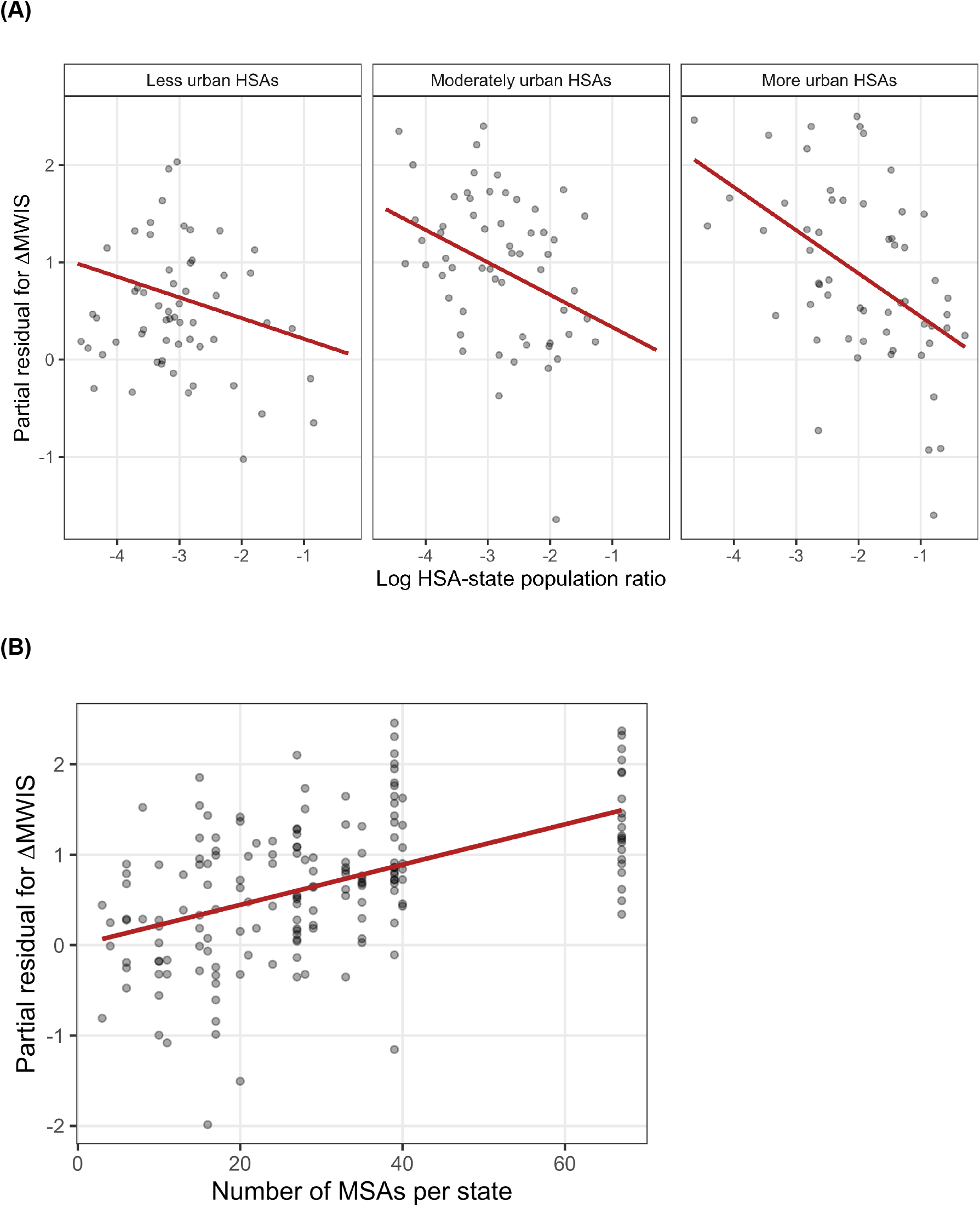
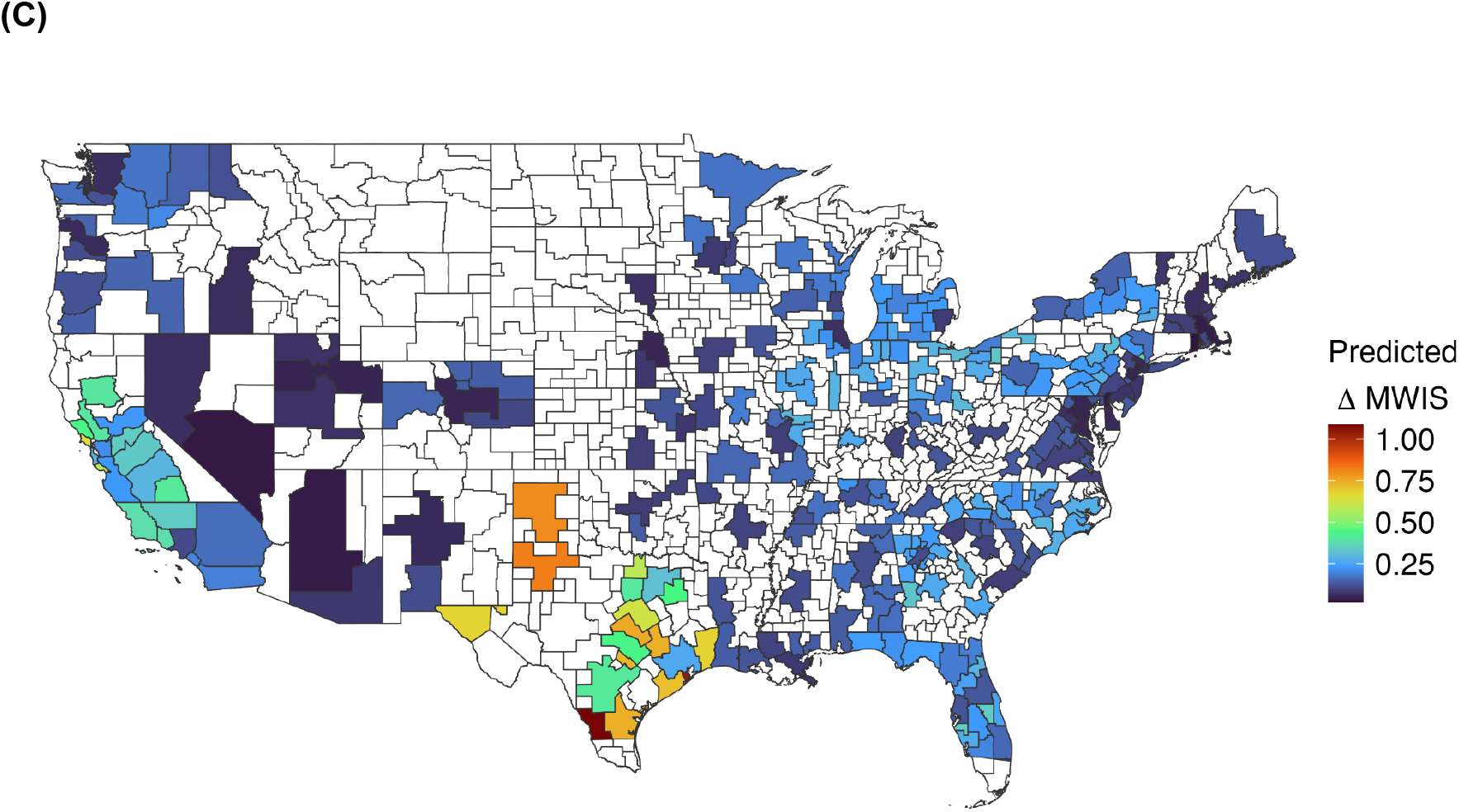
Population structure predicts the added value of local (HSA-level) forecasting over state-level models. GLM results relating one-week-ahead forecast improvement (ΔMWIS) to the HSA–state population ratio, urbanization, and the number of metropolitan statistical areas (MSAs) per state. (A) Partial residual plots stratified by terciles of HSA urbanization. Red lines show the predicted relationship at the median urbanization level within each tercile, holding other covariates constant. The negative association between HSA-state population and forecast improvement is strongest in more urbanized HSAs. (B) Partial residual plot showing the relationship between MSA count and forecast improvement after controlling for population ratio, urbanization, and their interaction. The positive slope (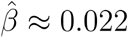) indicates positive gains from local forecasting in states with larger numbers of MSAs. (C) Projected one-week-ahead forecast improvement for HSAs with populations ≥250,000, based on the fitted GLM. Colors indicate the expected improvement of local forecasts relative to state-level forecasts, with warmer colors corresponding to larger gains. Because all mapped values are positive, even cooler colors indicate modest improvements from local forecasting. White regions indicate HSAs with populations below 250,000 that were excluded from the analysis. Values are back-transformed to the original scale. Connecticut HSAs were excluded because updated FIPS codes could not be matched to population density estimates obtained through tidycensus.

We next evaluated model fit and used the fitted GLM to project the value of local forecasting nationwide. Observed versus predicted 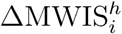 values for HSAs with complete data suggest that the model captures systematic variation in forecast improvement across regions (Supplementary Figure C.S1(A)). We then applied the fitted model to all HSAs with populations above 250,000 to estimate where local forecasting is expected to provide the greatest benefit. The resulting map reveals pronounced geographic variation in projected gains. The largest improvements are concentrated in highly fragmented, multi-metro states such as California and Texas, as well as parts of the Southeast. In contrast, more homogeneous and sparsely populated regions show relatively limited benefit from local forecasting (Figure 3C).

## Discussion

Our results demonstrate that local-level forecasts generally provide more accurate and reliable predictions of influenza activity than state-level forecasts in mid-sized and large local jurisdictions, especially at shorter horizons. Across all three evaluation metrics—coverage rate, mean absolute error (MAE), and mean weighted interval score (MWIS)—local models achieved better coverage and lower error in a majority of jurisdictions studied, indicating better-calibrated uncertainty and improved point accuracy. These gains reflect the ability of finer-scale models to capture local epidemic dynamics that are smoothed or obscured by state-level aggregation. Importantly, however, the advantage of local forecasting is not uniform across regions. While local forecasts reduced MWIS by 39.2% at the one-week horizon, this improvement declined to 19.6%, 11.4%, and 6.5% at two-, three-, and four-weeks ahead, indicating reduced gains and greater variability at longer horizons. Consistent with this pattern, the proportion of HSAs in which local forecasts outperformed state forecasts decreased from 98.8% at one week to 90.8%, 78.6%, and 69.4% at two-, three-, and four-week ahead. Regression analyses show that the largest improvements occurred in HSAs that make up a smaller share of their state’s population, particularly ones in more urban areas, and in states subdivided into a larger number of MSAs—settings in which epidemic timing and intensity vary more across regions within the same state.

These findings have direct implications for how models could be incorporated into public health practice. Many preparedness and response decisions—such as healthcare surge planning, targeted outreach, and risk communication—are made at local or regional levels rather than statewide. In these settings, forecasts that more accurately reflect local epidemic trajectories can provide more timely and actionable guidance. In our analysis, HSA-level forecasts were on average more accurate and better calibrated than state-level forecasts across horizons, suggesting that local forecasting in larger HSAs can in many settings better support operational decision-making. Our results suggest that investments in local forecasting capacity are most likely to pay off in large, heterogeneous states and populous metropolitan regions where epidemic dynamics diverge from statewide averages. However, we did not evaluate smaller or rural jurisdictions (populations <250,000), where greater stochastic variability may constrain forecastability. Accordingly, while our findings support the value of local forecasting in larger substate regions, further work is needed to assess performance in smaller or less populous settings.

A key feature of this study is the use of facility-level emergency department visit data from the National Syndromic Surveillance Program, aggregated to the HSA level. Although public health stakeholders often prioritize outcomes such as hospitalizations, case counts, or deaths, such measures are rarely available nationwide at fine spatial resolution and usually require additional normalization to enable fair comparison. Count-based outcomes scale with population size, and at local levels the appropriate denominator is the facility catchment population, which generally requires patient-level data that are difficult to obtain consistently across jurisdictions. By contrast, the percentage of ED visits attributable to influenza is publicly available for approximately 70% of HSAs and is intrinsically normalized, enabling meaningful comparisons of forecasting performance across regions without additional catchment adjustment. While the percentage of ED visits attributable to influenza is not a direct measure of disease burden, it provides a timely and spatially consistent indicator of local epidemic dynamics that is available nationwide at substate resolution.

This study has several limitations. First, our analysis focuses on HSAs with populations larger than 250,000 because smaller HSAs often exhibited noisier surveillance signals, likely due to lower patient volumes and greater variability in reported visit counts. Restricting the analysis to larger HSAs enabled more stable model fitting and evaluation but limits the generalizability of our findings to smaller regions. For diseases with higher hospitalization rates, smaller HSAs may generate sufficient data to support similar analyses. Accordingly, our results should be interpreted as applying primarily to seasonal influenza in mid- to large-sized HSAs.

Second, in modeling where local forecasts provide the greatest gains, we intentionally restricted predictors to a small set of interpretable population-structure variables. Our goal was not to maximize predictive accuracy, but to obtain transparent, generalizable insights into how regional characteristics relate to the value of local forecasting. More flexible machine-learning approaches could potentially improve predictive performance or capture higher-order interactions, but their results are often less interpretable. In contrast, the GLM framework provides clear effect estimates that summarize how key structural features are associated with differences in forecast accuracy.

Third, because the regression analyses were conducted at the location level after averaging MWIS over reference dates and fitting separate models by horizon, the dependence structure was reduced compared with using forecast-date-level observations directly. However, locations may still exhibit residual dependence due to shared spatial, seasonal, and surveillance-related structure. Therefore, regression-based inferences should be interpreted cautiously and statistical significance estimates should be interpreted with caution since standard error estimates may be under-estimated.

Finally, our comparison implicitly assumes that statewide forecasts are used to inform local decision-making by applying state predictions to substate regions. While this reflects common practice in settings when local forecasts are unavailable, it may not capture the full range of operational uses. For example, officials may recalibrate state forecasts to recent local data while preserving projected trends––an adaptation not captured in our framework. Thus, the local utility of statewide forecasts depends in part on how they are interpreted and applied in practice.

More broadly, our findings highlight the importance of matching forecasting resolution to the scale at which epidemic dynamics evolve. In the early months of the COVID-19 pandemic, local outbreaks often emerged asynchronously and followed idiosyncratic trajectories within the same state, limiting the usefulness of statewide indicators for anticipating local conditions. As surveillance systems expand and future epidemics may increasingly spread through heterogeneous urban networks, forecasting approaches that operate at local scales may provide earlier and more relevant signals of changing risk. Our results suggest that targeted investment in local forecasting—particularly in large and demographically diverse states—can improve situational awareness and support more timely and spatially informed public health decisions for both seasonal influenza and emerging infectious disease threats.

## Materials and Methods

### Data

We analyzed weekly estimates of the percentage of emergency department (ED) visits attributable to influenza from the CDC National Syndromic Surveillance Program (NSSP) (29). These publicly available data are derived from chief complaint text and discharge diagnosis fields using ICD-10-CM influenza codes and are aggregated by epidemiological week at the Health Service Area (HSA) level, a geography defined by hospital referral patterns. The dataset covers approximately 68% of US counties and 43.4% of the US population (Supplementary Fig. A.S7). We analyzed percentages rather than raw visit counts because percentages are intrinsically normalized across regions and avoid the need to estimate hospital catchment populations, which are difficult to define consistently across heterogeneous reporting systems. Percentage-based outcomes also reduce scale-related distortions in forecast evaluation metrics such as weighted interval score (WIS). Comparisons of influenza-related ED visit counts and percentage-based indicators showed highly similar temporal dynamics, supporting percentages as a stable measure of influenza activity.

### Forecasting Framework

All primary forecasts were generated using a Gradient Boosting Quantile Regression (GBQR) framework (30), which estimates conditional quantiles of future influenza activity given predictor variables *x*_*i,t*_. We additionally evaluated an automated ARIMA model and a quantile baseline model; details for these approaches are provided in the Supplementary Material D, with summary results shown in Supplementary Tables A.S1-A.S4.

For each forecast horizon *h* and quantile level *τ*, the model estimates

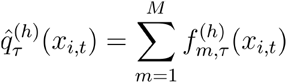

Where 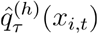 denotes the predicted *τ*-th conditional quantile of influenza activity at time *t* + *h*, and 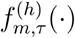 denotes regression trees added sequentially by gradient boosting.

Quantiles were estimated separately using pooled training data across all locations, and forecasts were ensembled across 100 random training subsamples to improve stability. Predictive distributions were summarized using the median forecast together with 95% prediction intervals. Models were implemented using the LightGBM quantile regression framework in Python (31).

### Retrospective Forecasting Design

We conducted retrospective forecasting experiments across the 2022–2023, 2023–2024, and 2024–2025 influenza seasons. For each target season, models were trained using observations from the remaining two seasons together with all observations available prior to the forecast date within the target season. Forecasts were generated weekly from October through March for 1–4 week horizons, with models re-fit at each forecast date. A single GBQR model was trained using pooled data across all included HSAs and states.

### Forecast Evaluation

We evaluated forecast performance using three standard metrics: 95% prediction interval coverage, mean absolute error (MAE), and weighted interval score (WIS) (20). Together, these capture point forecast accuracy, probabilistic calibration, and forecast sharpness. We summarize probabilistic forecast performance using mean weighted interval score (MWIS), computed by averaging WIS across forecast dates for each HSA and forecast horizon. Lower MAE and MWIS values indicate improved forecast performance. Formal definitions of all evaluation metrics are provided in Supplementary Material E.

To quantify differences in forecast performance at the HSA level, we compared the mean weighted interval score (MWIS) of HSA-level and state-level forecasts. Let *i* denote an HSA and *s*(*i*) the corresponding state. For forecast horizon *h*, we defined the average improvement in forecast performance as

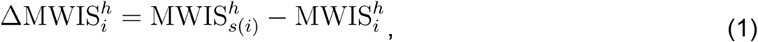

Where 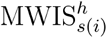 and 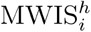 denote the performance of corresponding state-level and HSA-level forecasts, respectively, both evaluated against HSA-level observations. Positive values indicate improved performance of HSA-level forecasts relative to state-level forecasts.

### Modeling Predictors of Forecast Improvement

To identify settings in which local forecasting provides the greatest added value, we modeled forecast improvement as a function of regional population structure and urbanization, motivated by prior work linking these factors to influenza transmission dynamics and spatial heterogeneity (32, 33). Candidate predictors included measures of population size, urbanization, demographic composition, and spatial fragmentation. Final variable selection emphasized interpretability, stability across forecast horizons, and consistency across model specifications, with additional details provided in Supplementary Material C.

The final model included the HSA–state population ratio, urban population share, their interaction, and the number of Metropolitan Statistical Areas (MSAs) within each state. We hypothesized that MSA count would capture substate heterogeneity in epidemic timing because MSAs reflect major population centers and commuting structure. Population and metropolitan delineation data were obtained from the U.S. Census Bureau and Office of Management and Budget (34–36).

We evaluated generalized linear models (GLM), generalized additive models (GAM), and Bayesian additive regression trees (BART). Because all approaches produced qualitatively similar results, we report GLM results in the main text to prioritize interpretability; alternative specifications and diagnostics are provided in Supplementary Material C.

Let 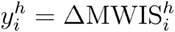 denote the average forecast improvement for HSA *i* at forecast horizon *h*. The final GLM was specified as

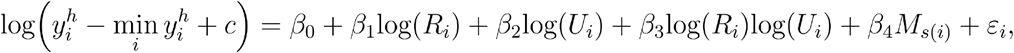

Where *R*_*i*_ denotes the HSA-state population ratio, *U*_*i*_ denotes the proportion of the HSA population residing in urban-designated areas, and *M*_*s*(*i*)_ denotes the number of MSAs in the corresponding state. Models were fit separately for each horizon.

## Data Availability

All data produced are available online at https://data.cdc.gov/Public-Health-Surveillance/NSSP-Emergency-Department-Visit-Trajectories-by-St/rdmq-nq56/about_data

## Acknowledgments

This work was supported by grants from the Centers for Disease Control and Prevention (U01IP001136, NU38FT000008, and 75D30122C14776), the Council of State and Territorial Epidemiologists (NU38OT000297), the National Science Foundation (2230125), and the National Institutes of General Medical Sciences (R35GM119582). The content is solely the responsibility of the authors and does not necessarily represent the official views of the funding agencies.

## Author Contributions

D.K. and L.A.M. designed research; D.K. performed research; D.K. analyzed data; D.K., R.P., K.E.J., S.J.F., N.G.R.,and L.A.M.wrote the paper.

