## Supplementary material for "Local Influenza Forecasts Outperform State-Level Forecasts in the United States"

### Supplementary Material A — Additional Figures and Tables

**(A)**

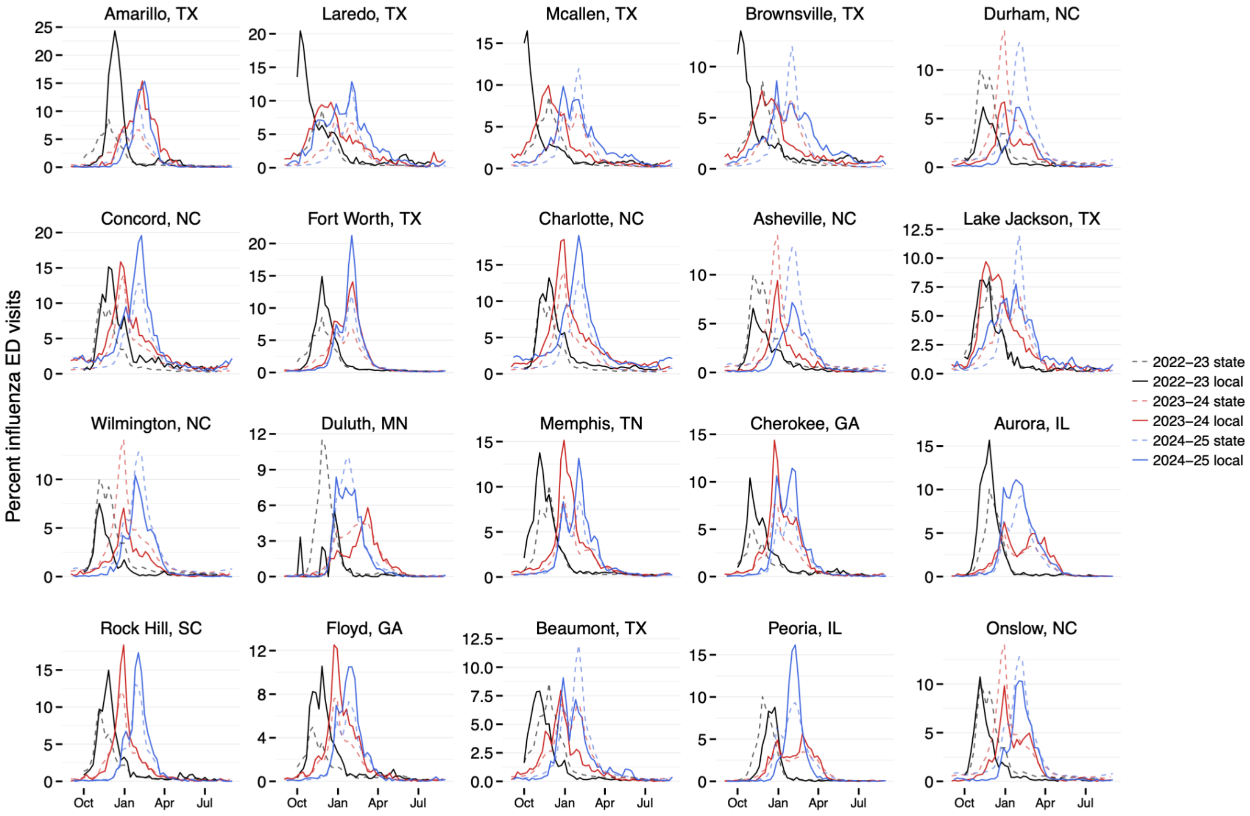


**(B)**

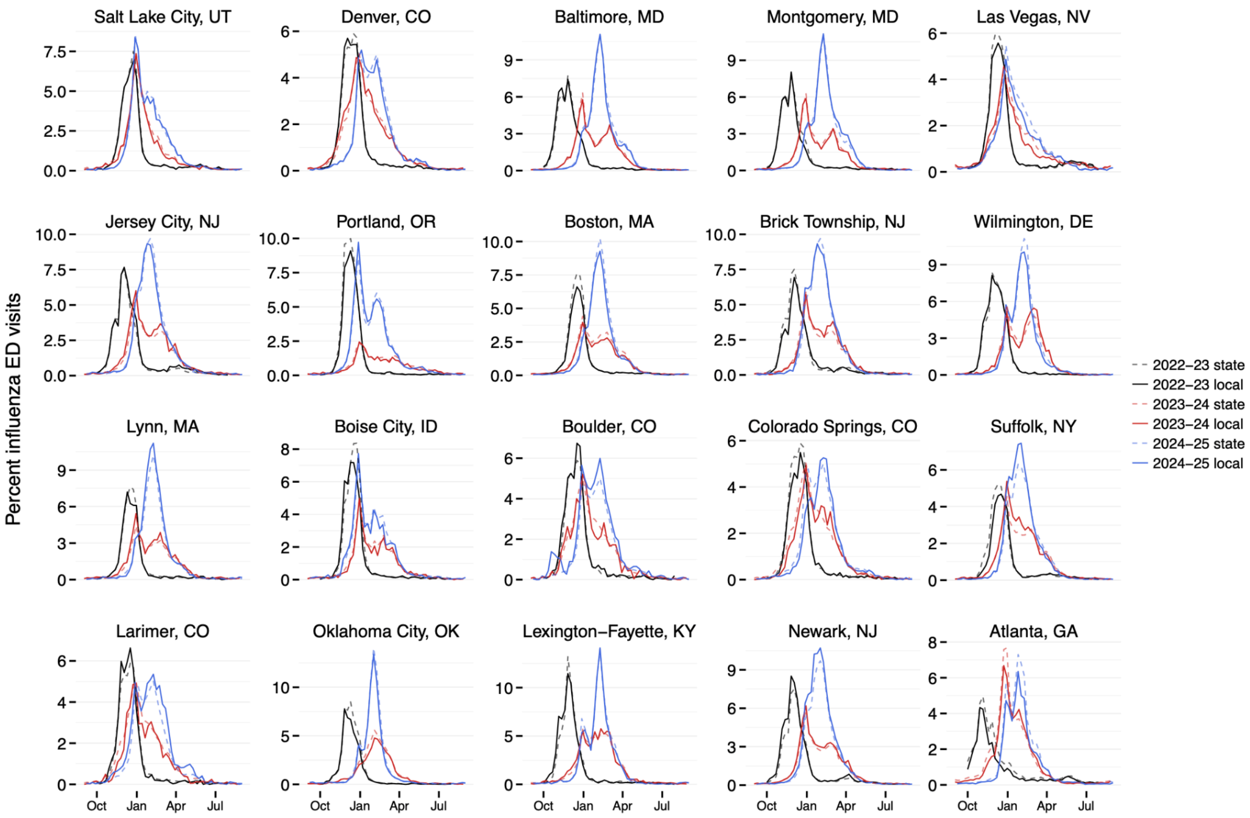


**Fig.A.S1. Comparison of HSA-level and state-level trends in the percentage of emergency department (ED) visits attributable to influenza during the 2022–2023, 2023–2024, and 2024–2025 influenza seasons.** (A) Twenty HSAs selected as extreme examples illustrate large discrepancies between HSA- and state-level influenza trends. These HSAs correspond to the 20 regions with the highest overall root mean squared error (RMSE) values (Table A.S2). Panels are arranged in descending order of overall RMSE. (B) Twenty HSAs selected as extreme examples illustrate minimal discrepancies between HSA- and state-level influenza trends. These HSAs correspond to the 20 regions with the lowest overall RMSE values (Table A.S2). Panels are arranged in ascending order of overall RMSE.

**(A)**

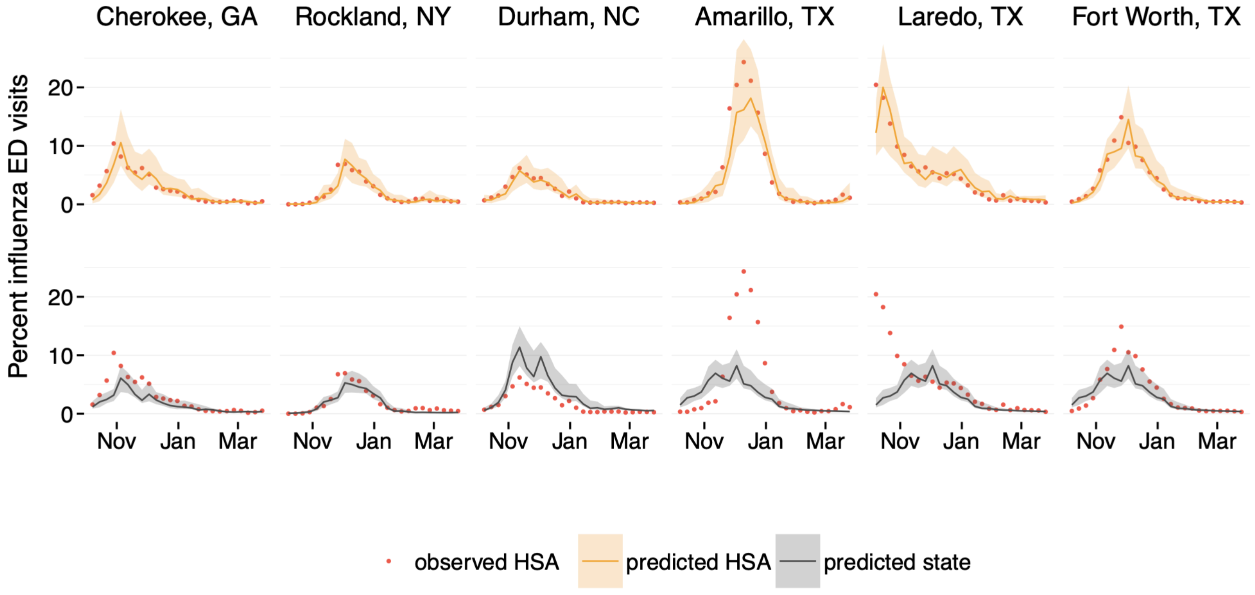

Cherokee, GA
Rockland, NY
Durham, NC
Amarillo, TX
Laredo, TX
Fort Worth, TX
Percent influenza ED visits
0
10
20
Nov
Jan
Mar
observed HSA
predicted HSA
predicted state


**(B)**

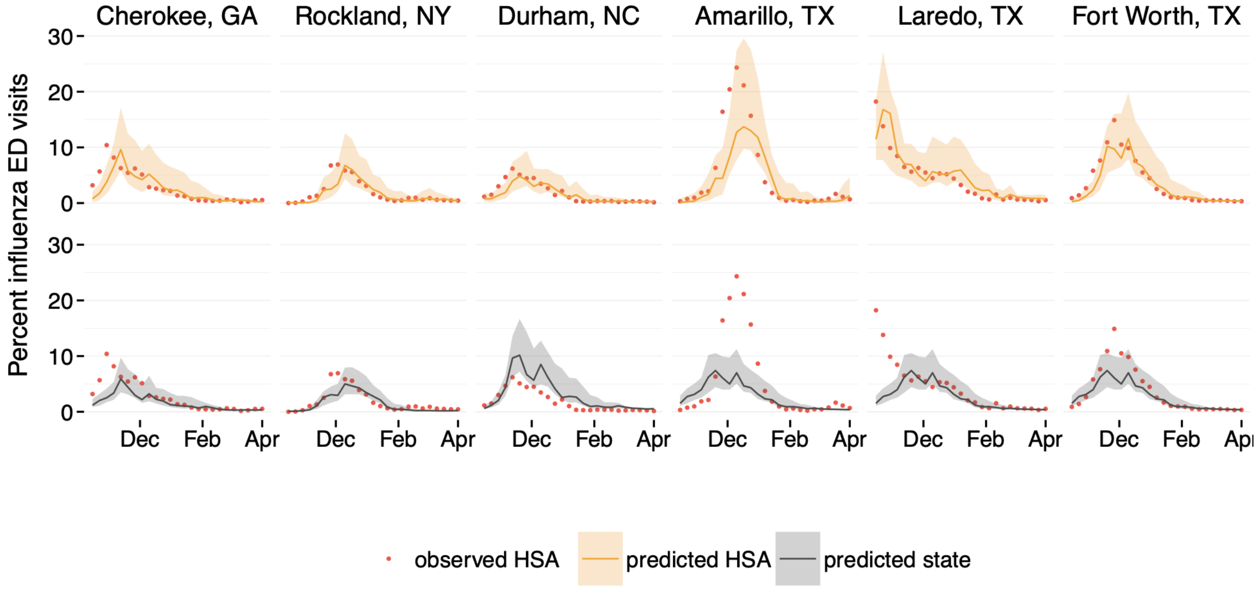

Cherokee, GA
Rockland, NY
Durham, NC
Amarillo, TX
Laredo, TX
Fort Worth, TX
Percent influenza ED visits
0
10
20
30
Dec
Feb
Apr
observed HSA
predicted HSA
predicted state

**(C)**

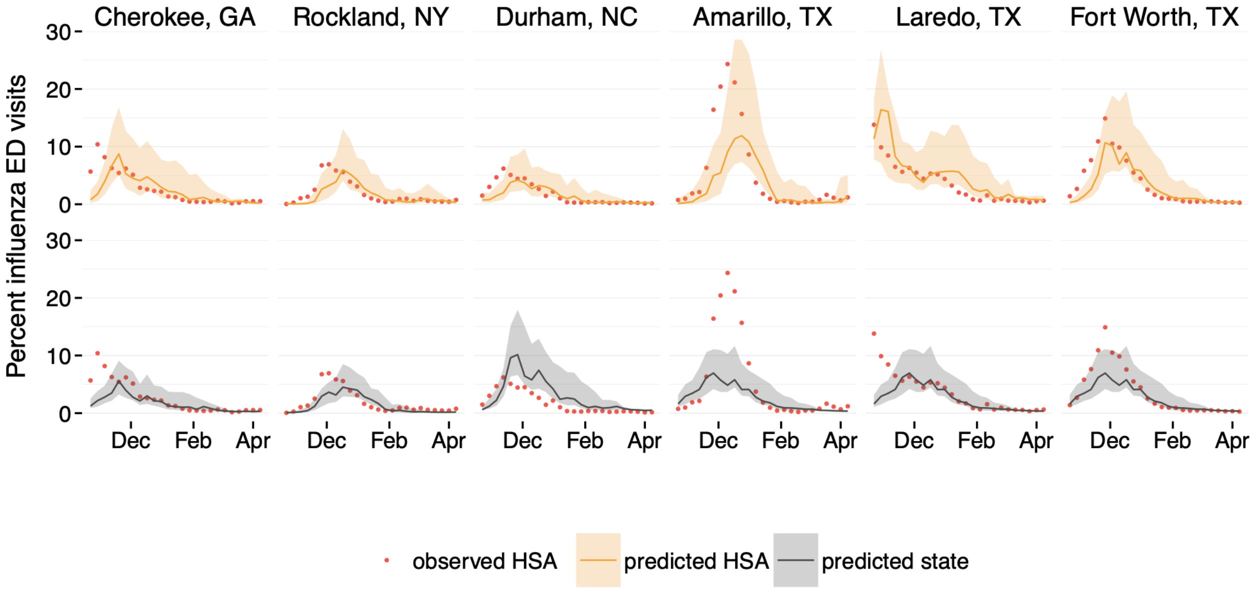


**(D)**

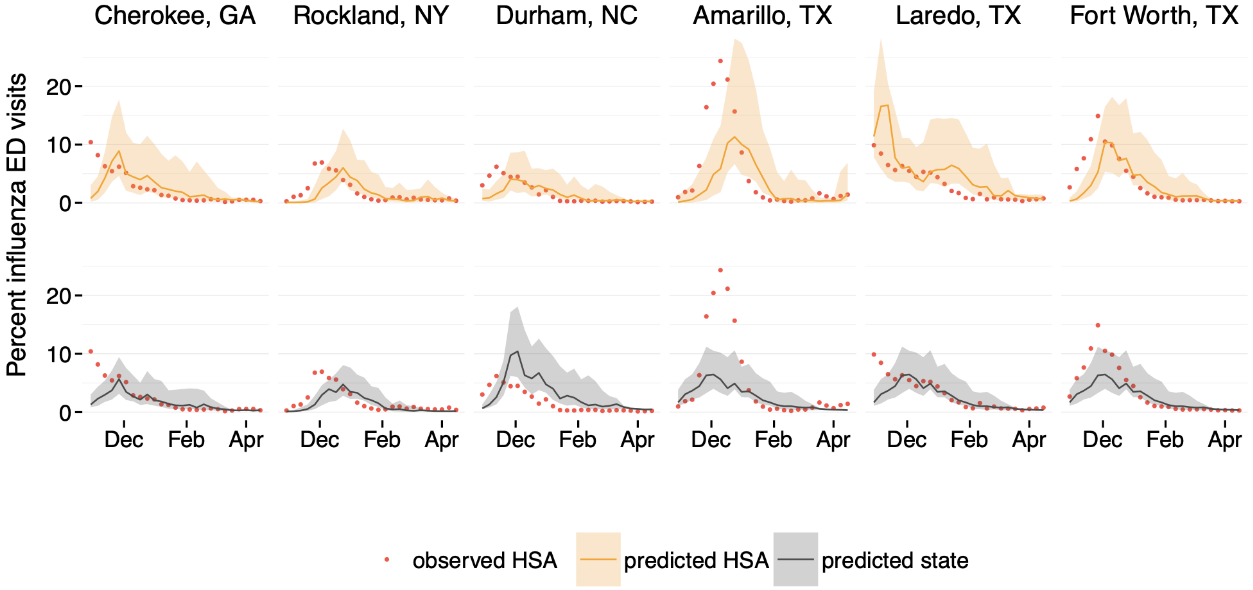


**Fig. A.S2. Forecasts of the percentage of emergency department (ED) visits attributable to influenza for six metropolitan Health Service Areas (HSAs) during the 2022–2023 influenza season.** Red points indicate observed HSA-level ED visit percentages. Solid lines represent forecast medians, and shaded regions indicate 95% prediction intervals. (A–D) Daily fixed-horizon forecasts concatenated over time across successive forecast initiation dates, corresponding to one-, two-, three-, and four-week-ahead horizons, respectively. In each panel, the top row shows forecasts generated using HSA-level data (orange), and the bottom row shows forecasts generated using state-level data (gray), each compared to observed HSA-level ED visits.

(A)

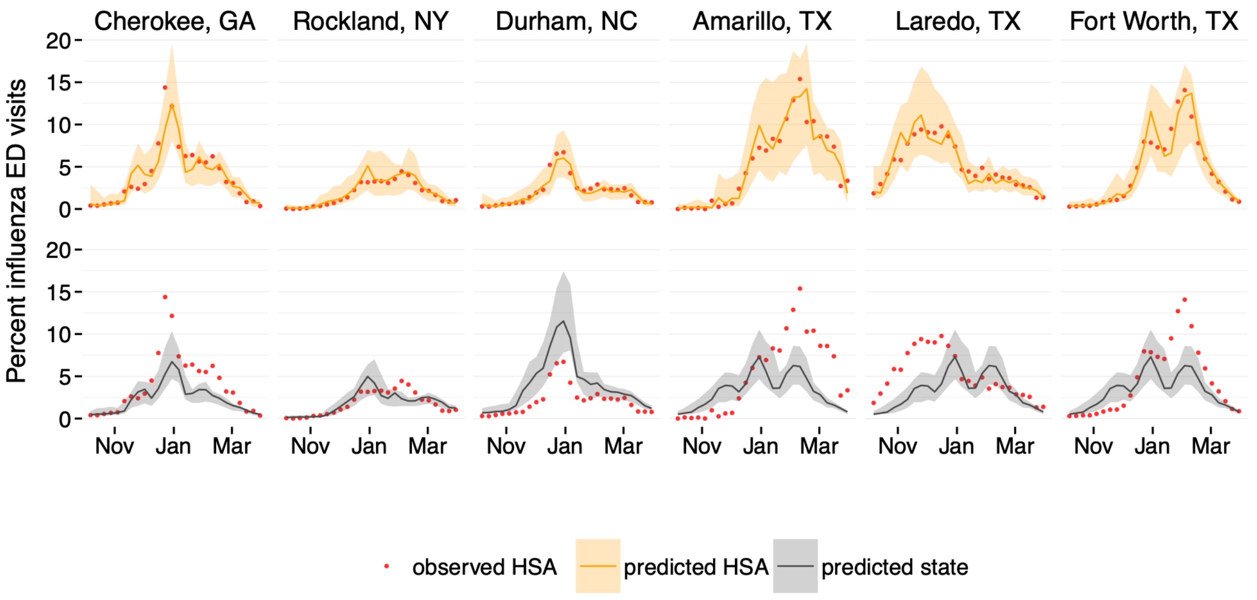

Cherokee, GA
Rockland, NY
Durham, NC
Amarillo, TX
Laredo, TX
Fort Worth, TX
Percent influenza ED visits
0
5
10
15
20
Nov
Jan
Mar
observed HSA
predicted HSA
predicted state


(B)

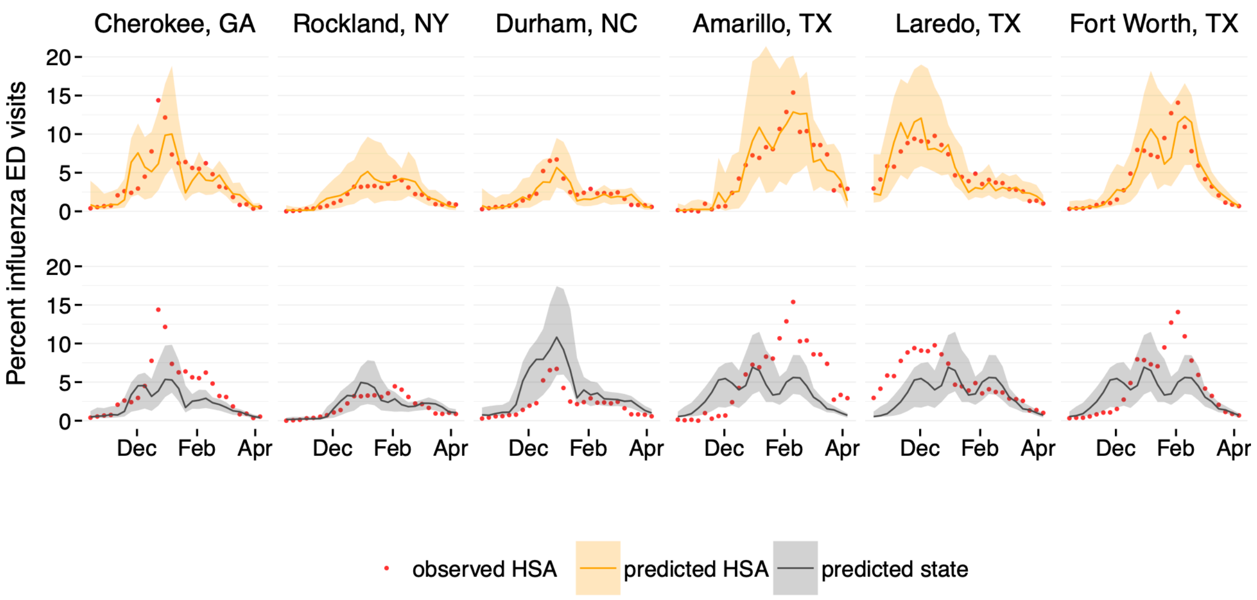

Cherokee, GA
Rockland, NY
Durham, NC
Amarillo, TX
Laredo, TX
Fort Worth, TX
Percent influenza ED visits
0
5
10
15
20
Dec
Feb
Apr
observed HSA
predicted HSA
predicted state

**(C)**

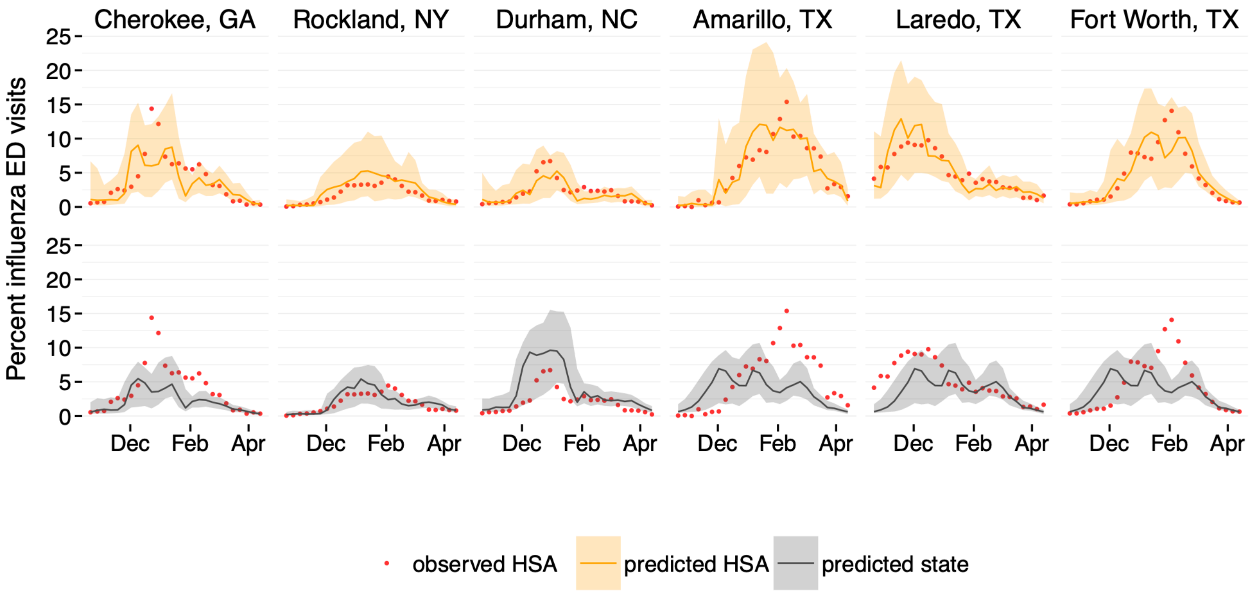


**(D)**

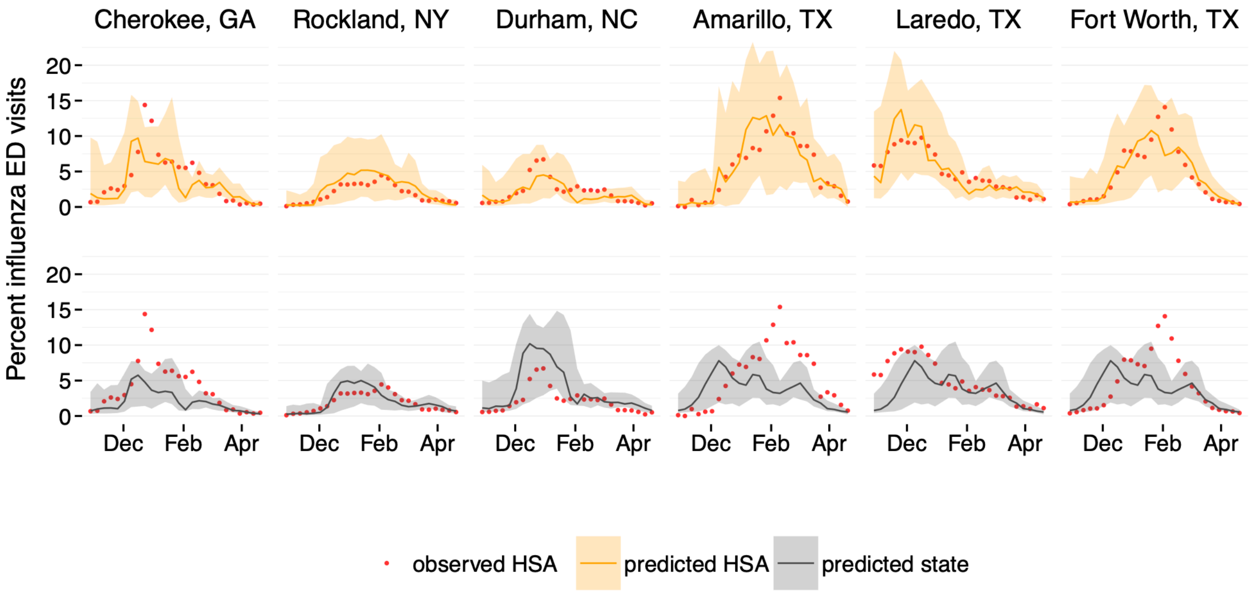


**Fig. A.S3. Forecasts of the percentage of emergency department (ED) visits attributable to influenza for six metropolitan Health Service Areas (HSAs) during the 2023–2024 influenza season.** Red points indicate observed HSA-level ED visit percentages. Solid lines represent forecast medians, and shaded regions indicate 95% prediction intervals. (A–D) Daily fixed-horizon forecasts concatenated over time across successive forecast initiation dates, corresponding to one-, two-, three-, and four-week-ahead horizons, respectively. In each panel, the top row shows forecasts generated using HSA-level data (orange), and the bottom row shows forecasts generated using state-level data (gray), each compared to observed HSA-level ED visits.

**(A)**

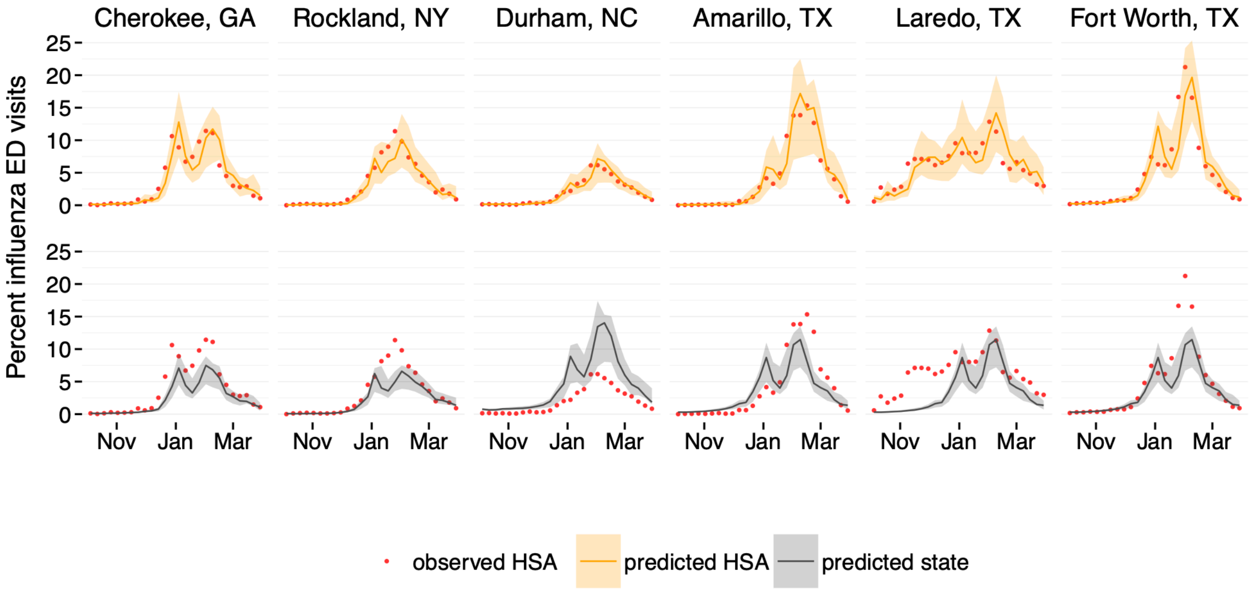
Cherokee, GA
Rockland, NY
Durham, NC
Amarillo, TX
Laredo, TX
Fort Worth, TX
Percent influenza ED visits
0
5
10
15
20
25
Nov
Jan
Mar
observed HSA
predicted HSA
predicted state

**(B)**

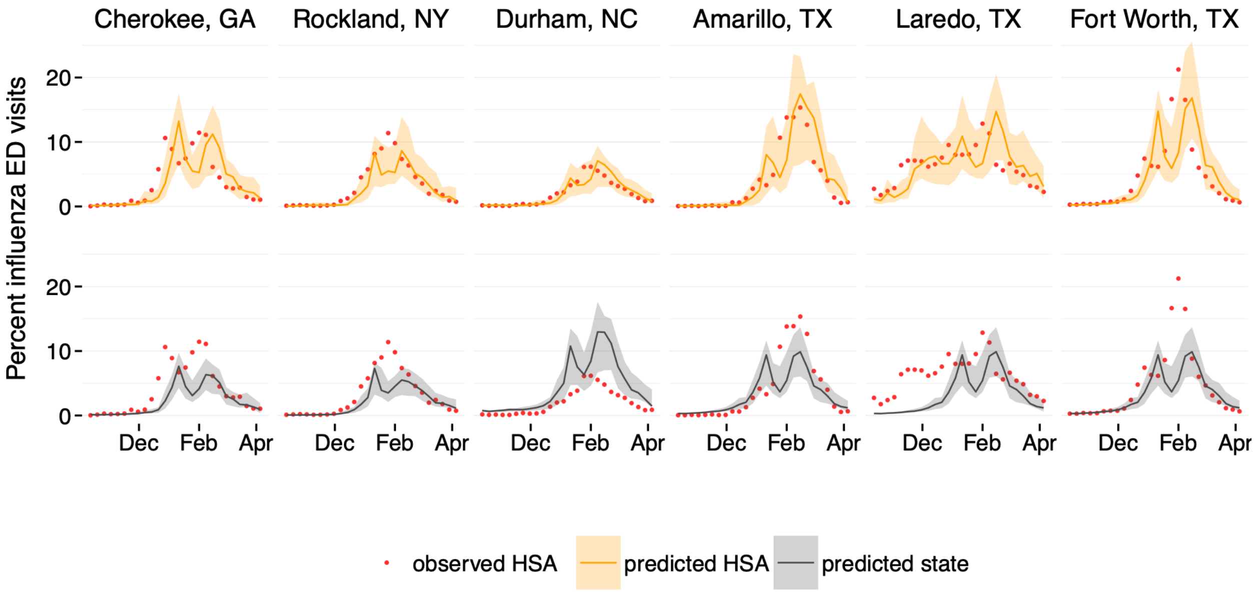
Cherokee, GA
Rockland, NY
Durham, NC
Amarillo, TX
Laredo, TX
Fort Worth, TX
Percent influenza ED visits
0
10
20
Dec
Feb
Apr
observed HSA
predicted HSA
predicted state

(C)

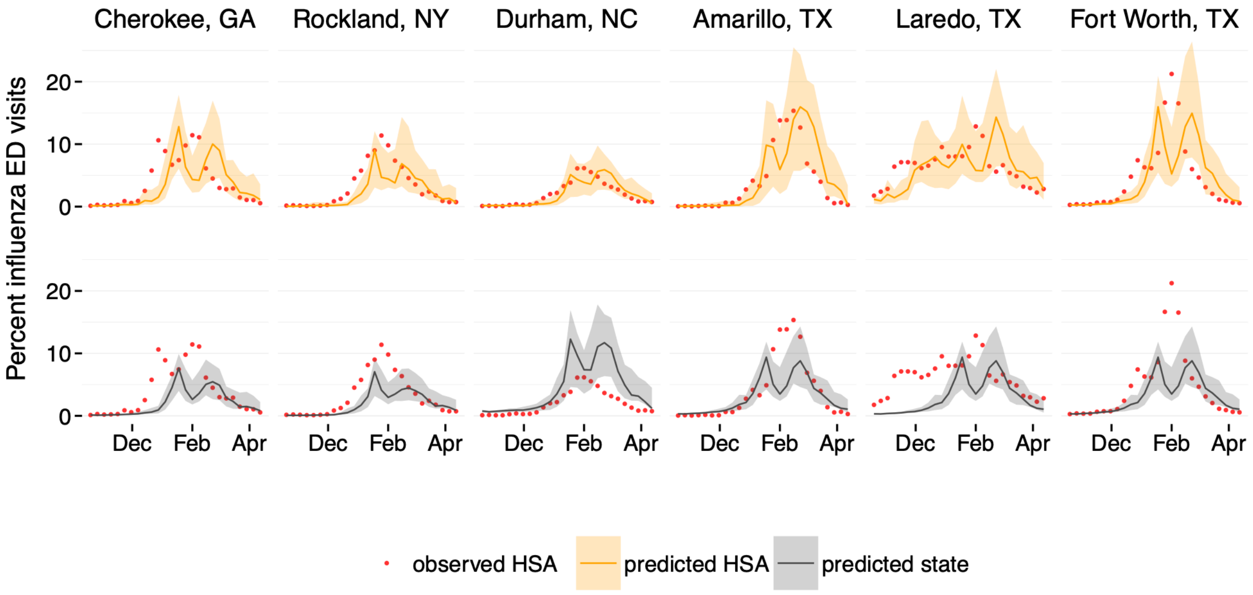


(D)

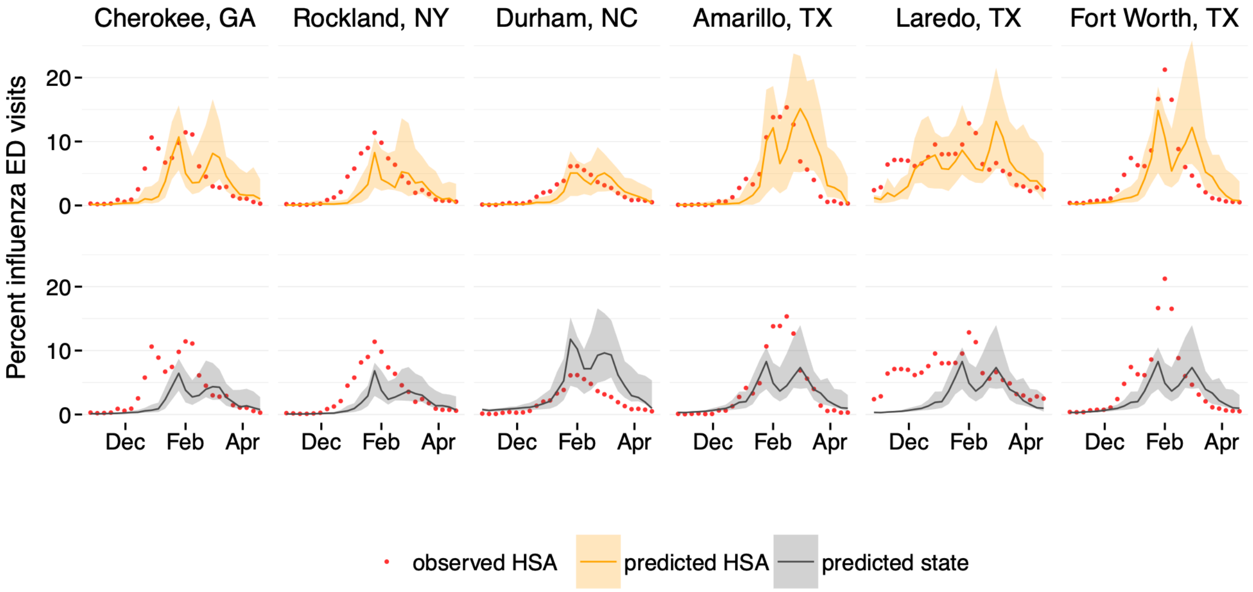


**Fig. A.S4. Forecasts of the percentage of emergency department (ED) visits attributable to influenza for six metropolitan Health Service Areas (HSAs) during the 2024–2025 influenza season.** Red points indicate observed HSA-level ED visit percentages. Solid lines represent forecast medians, and shaded regions indicate 95% prediction intervals. (A–D) Daily fixed-horizon forecasts concatenated over time across successive forecast initiation dates, corresponding to one-, two-, three-, and four-week-ahead horizons, respectively. In each panel, the top row shows forecasts generated using HSA-level data (orange), and the bottom row shows forecasts generated using state-level data (gray), each compared to observed HSA-level ED visits.

**(A)**

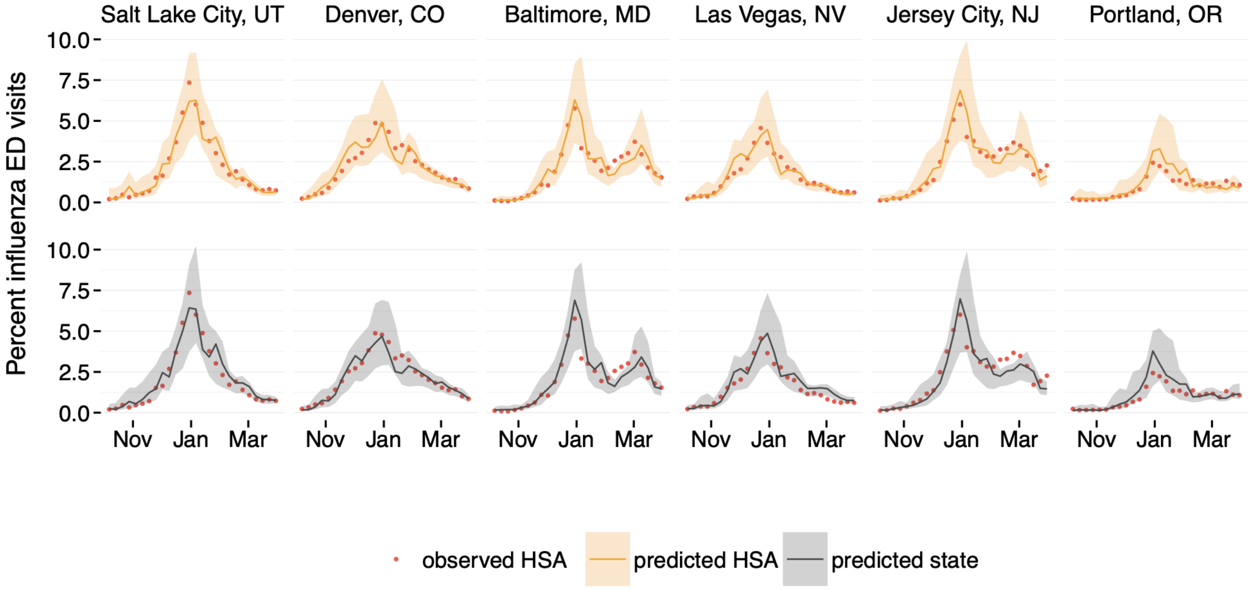
Salt Lake City, UT
Denver, CO
Baltimore, MD
Las Vegas, NV
Jersey City, NJ
Portland, OR
Percent influenza ED visits
10.0
7.5
5.0
2.5
0.0
Nov
Jan
Mar
observed HSA
predicted HSA
predicted state

**(B)**

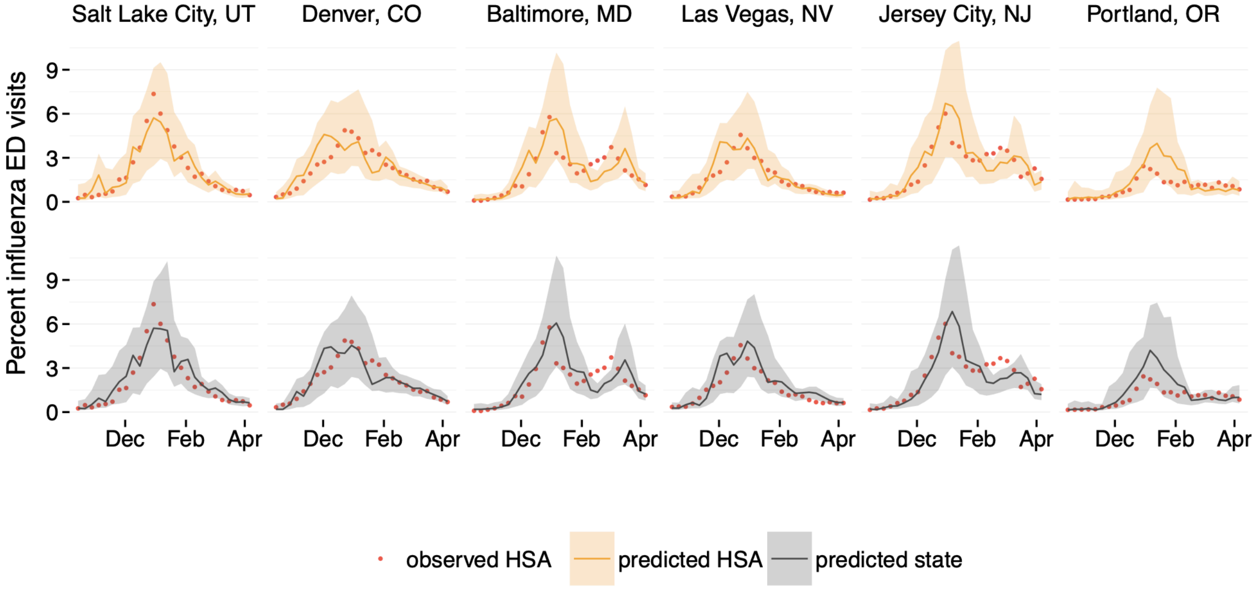
Salt Lake City, UT
Denver, CO
Baltimore, MD
Las Vegas, NV
Jersey City, NJ
Portland, OR
Percent influenza ED visits
9
6
3
0
Dec
Feb
Apr
observed HSA
predicted HSA
predicted state

(C)

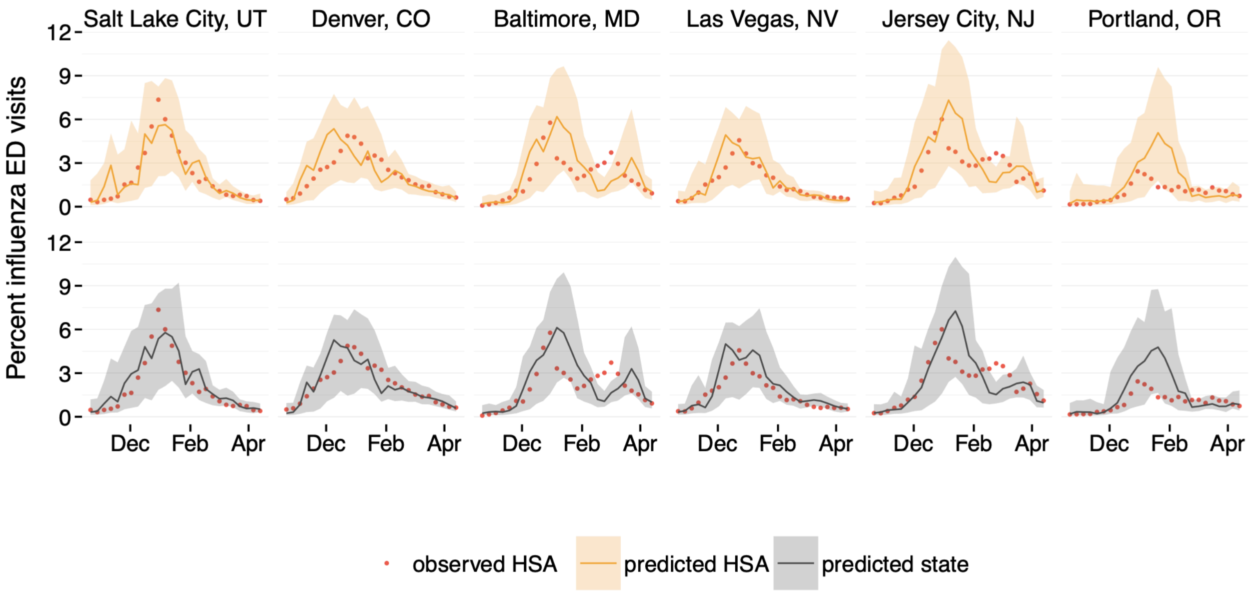


(D)

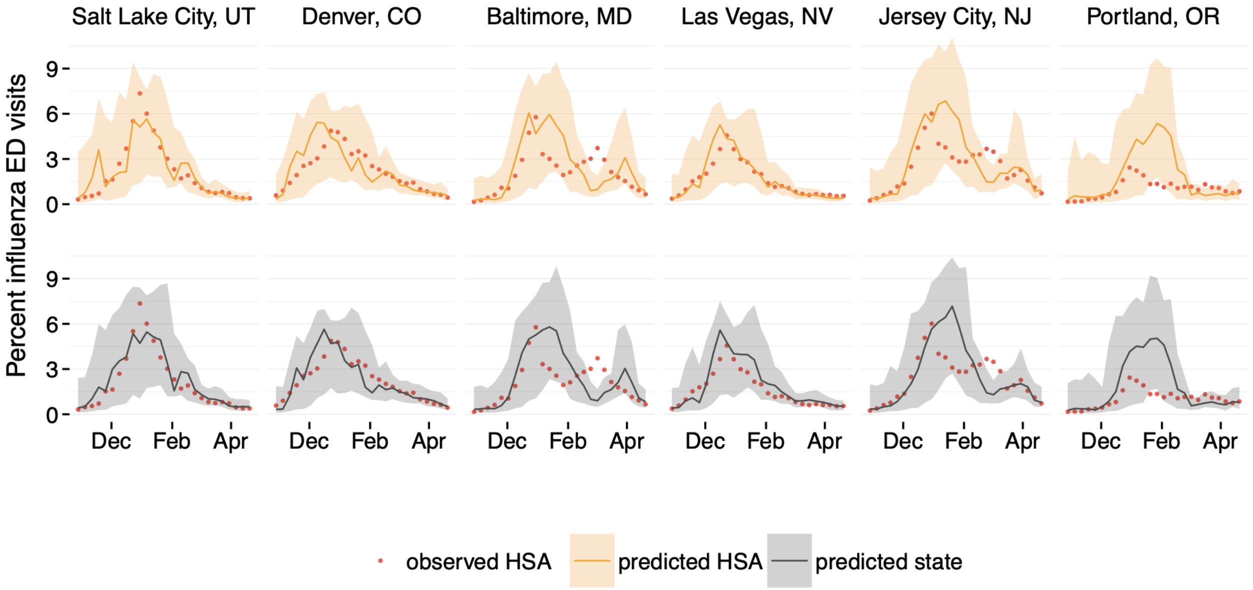


**Fig. A.S5. Forecasts of the percentage of emergency department (ED) visits attributable to influenza for six metropolitan Health Service Areas (HSAs) with small HSA–state discrepancies during the 2023–2024 influenza season.** Red points indicate observed HSA-level ED visit percentages. Solid lines represent forecast medians, and shaded regions indicate 95% prediction intervals. (A–D) Daily fixed-horizon forecasts concatenated over time across successive forecast initiation dates, corresponding to one-, two-, three-, and four-week-ahead horizons, respectively. In each panel, the top row shows forecasts generated using HSA-level data (orange), and the bottom row shows forecasts generated using state-level data (gray), each compared to observed HSA-level ED visits.

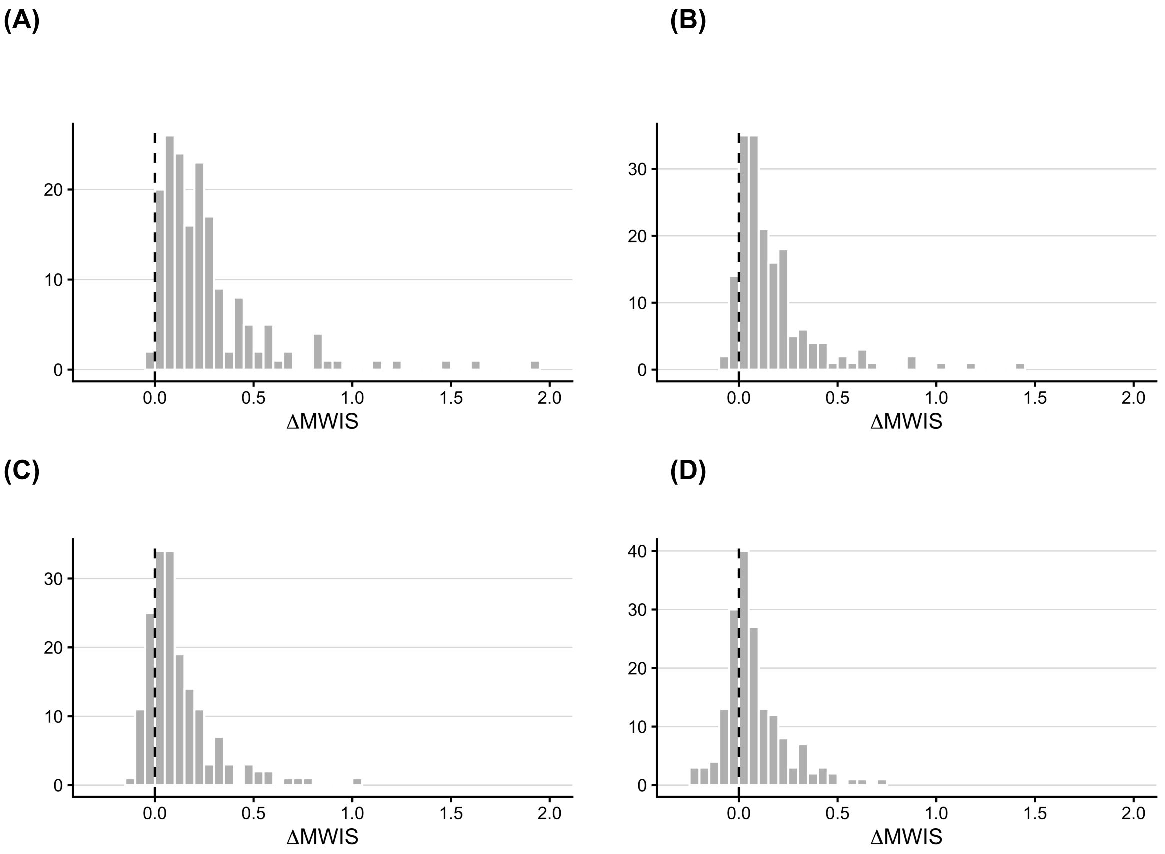


**Fig. A.S6**. **Histograms of HSA-specific average differences in Weighted Interval Score ($\Delta\mathrm{MWIS}$) between HSA-level and state-level forecasts for different horizons.** Panels A-D correspond to 1-2-3-, and 4-week-ahead forecast horizons, respectively. Positive values indicate improved performance at the HSA-level relative to the state-level aggregate across the 2022-2023, 2023-2024, and 2024-2025 seasons. The vertical dashed line at 0 denotes no difference in MWIS between HSA-level and state-level forecasts. For each HSA and forecast horizon, the average MWIS was first computed separately for the HSA-level and state-level forecasts by averaging across all forecast weeks and seasons. The plotted value represents the difference between these averages (MWIS_hsa - MWIS_state), resulting in one entry per HSA in each histogram.

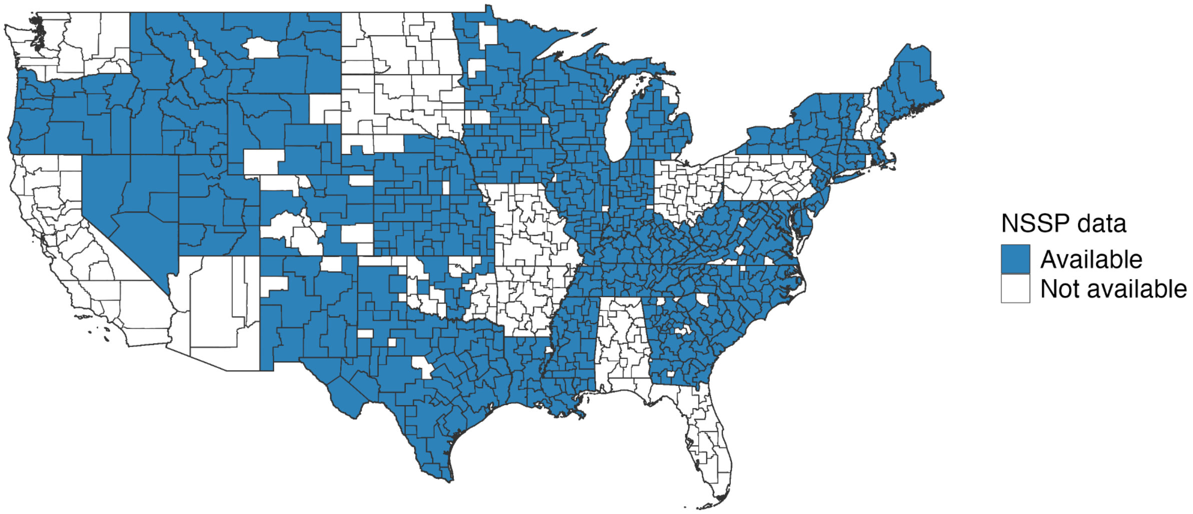


**Fig. A.S7**. **Data availability of the percentage of emergency department (ED) visits attributable to influenza by Health Service Area (HSA).** HSAs shaded in blue indicate those for which sub-state influenza trends are publicly available through the CDC HealthData.gov platform. Availability varies by state and region and reflects CDC criteria and jurisdiction-specific factors governing the public display of sub-state syndromic surveillance data.

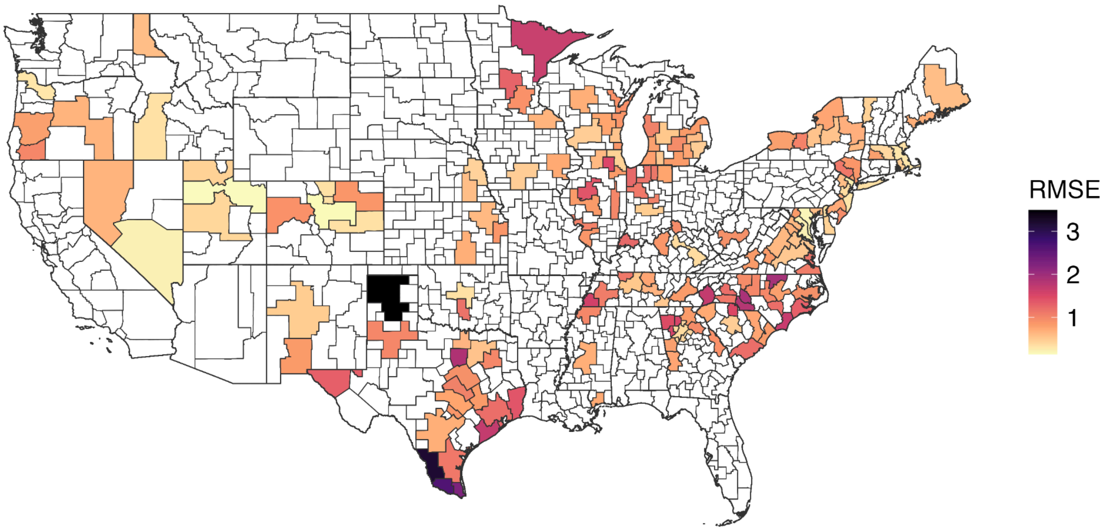


**Fig. A.S8. Health Service Areas (HSAs) included in the forecasting analysis.** Only HSAs with populations of at least 250,000 were included. Colors indicate overall root mean squared error (RMSE) values quantifying differences between HSA-level to state-level trends between October 2022 and July 2025 Table A.S5), with darker shades corresponding to larger discrepancies.

**Table A.S1. Average forecast accuracy of HSA-level versus state-level models across three influenza seasons (2022–2023, 2023–2024, and 2024–2025), for three different forecasting methods: GBQR, ARIMA, and baseline model that forecasts no change from the preceding week.** Performance is summarized for one-, two-, three-, and four-week-ahead forecast horizons. Metrics include coverage rate, mean absolute percentage error (MAE), and mean weighted interval score (MWIS), each averaged across all locations and forecast weeks. Coverage rates closer to the nominal 95% level indicate better-calibrated uncertainty, while lower MAE and MWIS values indicate greater forecast accuracy. Black bolded values indicate better performance between HSA- and state-level forecasts within a given model; red bolded values indicate the best performing model across all models and levels. Values in parentheses indicate standard deviations across locations and forecast weeks.

| Metrics | Model | Level | Forecast horizon | | | |
|---|---|---|---|---|---|---|
| | | | 1 | 2 | 3 | 4 |
| Coverage Rate | GBQR | HSA | **0.905 (0.293)** | **0.833 (0.373)** | **0.781 (0.413)** | **0.720 (0.449)** |
| | | State | 0.590 (0.492) | 0.614 (0.487) | 0.608 (0.488) | 0.580 (0.494) |
| | ARIMA | HSA | **0.853 (0.354)** | **0.849 (0.358)** | **0.842 (0.365)** | **0.839 (0.368)** |
| | | State | 0.741 (0.438) | 0.810 (0.392) | 0.823 (0.382) | 0.822 (0.382) |
| | BASELINE | HSA | **0.893 (0.310)** | **0.838 (0.369)** | **0.800 (0.400)** | **0.774 (0.418)** |
| | | State | 0.822 (0.382) | 0.799 (0.401) | 0.765 (0.424) | 0.738 (0.440) |
| MAE | GBQR | HSA | **0.599 (0.851)** | **0.981 (1.356)** | **1.254 (1.622)** | **1.388 (1.744)** |
| | | State | 0.908 (1.284) | 1.147 (1.556) | 1.349 (1.726) | 1.439 (1.774) |
| | ARIMA | HSA | **0.691 (0.923)** | **1.185 (1.583)** | 1.580 (2.006) | 1.876 (2.242) |
| | | State | 0.932 (1.311) | 1.268 (1.693) | **1.577 (2.000)** | **1.838 (2.194)** |
| | BASELINE | HSA | **0.760 (1.010)** | **1.313 (1.683)** | 1.789 (2.146) | 2.194 (2.465) |
| | | State | 0.993 (1.364) | 1.392 (1.770) | **1.787 (2.103)** | **2.153 (2.349)** |
| MWIS | GBQR | HSA | **0.300 (0.443)** | **0.525 (0.817)** | **0.705 (1.079)** | **0.852 (1.281)** |
| | | State | 0.567 (0.959) | 0.692 (1.104) | 0.821 (1.255) | 0.929 (1.384) |
| | ARIMA | HSA | **0.416 (0.669)** | **0.712 (1.153)** | **0.927 (1.458)** | **1.072 (1.619)** |
| | | State | 0.636 (1.137) | 0.798 (1.344) | 0.958 (1.533) | 1.082 (1.652) |
| | BASELINE | HSA | **0.471 (0.734)** | **0.813 (1.250)** | **1.098 (1.609)** | **1.330 (1.843)** |
| | | State | 0.672 (1.141) | 0.904 (1.417) | 1.142 (1.664) | 1.356 (1.848) |

**Table A.S2**. **Average forecast accuracy of HSA-level versus state-level models for three different influenza seasons (2022–2023, 2023–2024, and 2024–2025) using GBQR.** Performance is summarized for one-, two-, three-, and four-week-ahead forecast horizons. Metrics include coverage rate, mean absolute percentage error (MAE), and mean weighted interval score (MWIS), each averaged across all locations and forecast weeks, within a season. Coverage rates closer to the nominal 95% level indicate better-calibrated uncertainty, while lower MAE and MWIS values indicate greater forecast accuracy. Bold values denote superior performance of HSA-level forecasts. Values in parentheses indicate standard deviations across locations and forecast weeks.

| Season | Level | Forecast horizon | | | |
|---|---|---|---|---|---|
| | | 1 | 2 | 3 | 4 |
| 2022/23 | | **Coverage Rate** | | | |
| | HSA | **0.871 (0.336)** | **0.773 (0.419)** | **0.703 (0.457)** | **0.629 (0.483)** |
| | State | 0.543 (0.498) | 0.544 (0.498) | 0.508 (0.500) | 0.457 (0.498) |
| | | **MAE** | | | |
| | HSA | **0.571 (0.905)** | **0.955 (1.394)** | **1.197 (1.684)** | **1.438 (2.011)** |
| | State | 0.885 (1.457) | 1.082 (1.608) | 1.233 (1.708) | 1.409 (1.899) |
| | | **MWIS** | | | |
| | HSA | **0.298 (0.512)** | **0.525 (0.853)** | **0.724 (1.187)** | **0.950 (1.551)** |
| | State | 0.591 (1.171) | 0.691 (1.208) | 0.816 (1.323) | 0.988 (1.547) |
| 2023/24 | | **Coverage Rate** | | | |
| | HSA | **0.944 (0.229)** | **0.943 (0.233)** | **0.935 (0.246)** | **0.935 (0.247)** |
| | State | 0.647 (0.478) | 0.744 (0.437) | 0.793 (0.405) | 0.834 (0.372) |
| | | **MAE** | | | |
| | HSA | **0.518 (0.620)** | **0.783 (0.911)** | **1.005 (1.116)** | **1.079 (1.173)** |
| | State | 0.852 (1.056) | 1.026 (1.214) | 1.200 (1.371) | 1.237(1.384) |
| | | **MWIS** | | | |
| | HSA | **0.251 (0.271)** | **0.379 (0.413)** | **0.483 (0.520)** | **0.535 (0.559)** |
| | State | 0.512 (0.759) | 0.556 (0.763) | 0.614 (0.816) | 0.627 (0.822) |
| 2024/25 | | **Coverage Rate** | | | |
| | HSA | **0.900 (0.300)** | **0.783 (0.413)** | **0.703 (0.457)** | **0.594 (0.491)** |
| | State | 0.578 (0.494) | 0.551 (0.497) | 0.520 (0.500) | 0.445 (0.497) |
| | | **MAE** | | | |
| | HSA | **0.706 (0.976)** | **1.204 (1.632)** | **1.556 (1.914)** | **1.648 (1.892)** |
| | State | 0.987 (1.312) | 1.331 (1.778) | 1.610 (2.009) | 1.669 (1.959) |
| | | **MWIS** | | | |
| | HSA | **0.351 (0.501)** | **0.671 (1.032)** | **0.909 (1.315)** | **1.076 (1.442)** |
| | State | 0.598 (0.909) | 0.830 (1.258) | 1.033 (1.493) | 1.174 (1.594) |

**Table A.S3**. **Average forecast accuracy of HSA-level versus state-level models for three different influenza seasons (2022–2023, 2023–2024, and 2024–2025) using automated ARIMA.** Performance is summarized for one-, two-, three-, and four-week-ahead forecast horizons. Metrics include coverage rate, mean absolute percentage error (MAE), and mean weighted interval score (MWIS), each averaged across all locations and forecast weeks, within a season. Coverage rates closer to the nominal 95% level indicate better-calibrated uncertainty, while lower MAE and MWIS values indicate greater forecast accuracy. Bold values denote superior performance of HSA-level forecasts. Values in parentheses indicate standard deviations across locations and forecast weeks.

| Season | Level | Forecast horizon | | | |
|---|---|---|---|---|---|
| | | 1 | 2 | 3 | 4 |
| 2022/23 | | **Coverage Rate** | | | |
| | HSA | **0.847 (0.360)** | **0.826 (0.379)** | **0.811 (0.391)** | **0.806 (0.396)** |
| | State | 0.742 (0.437) | 0.788 (0.409) | 0.800 (0.400) | 0.791 (0.407) |
| | | **MAE** | | | |
| | HSA | **0.712 (1.006)** | **1.249 (1.677)** | 1.696 (2.141) | 2.048 (2.447) |
| | State | 0.978 (1.535) | 1.337 (1.863) | **1.675 (2.123)** | **1.991 (2.355)** |
| | | **MWIS** | | | |
| | HSA | **0.444 (0.750)** | **0.765 (1.229)** | **1.016 (1.575)** | 1.206 (1.812) |
| | State | 0.703 (1.368) | 0.870 (1.503) | 1.036 (1.634) | **1.197 (1.793)** |
| 2023/24 | | **Coverage Rate** | | | |
| | HSA | **0.908 (0.289)** | **0.920 (0.271)** | **0.919 (0.273)** | **0.927 (0.261)** |
| | State | 0.771 (0.420) | 0.873 (0.332) | 0.892 (0.310) | 0.904 (0.294) |
| | | **MAE** | | | |
| | HSA | **0.592 (0.740)** | **0.968 (1.251)** | **1.262 (1.580)** | **1.465 (1.739)** |
| | State | 0.852 (1.104) | 1.083 (1.391) | 1.312 (1.643) | 1.482(1.774) |
| | | **MWIS** | | | |
| | HSA | **0.329 (0.492)** | **0.539 (0.824)** | **0.683 (1.040)** | **0.763 (1.125)** |
| | State | 0.542 (0.921) | 0.628 (1.035) | 0.737 (1.173) | 0.801 (1.231) |
| 2024/25 | | **Coverage Rate** | | | |
| | HSA | **0.804 (0.397)** | **0.799 (0.400)** | **0.794 (0.404)** | **0.782 (0.413)** |
| | State | 0.709 (0.454) | 0.768 (0.422) | 0.776 (0.417) | 0.770 (0.421) |
| | | **MAE** | | | |
| | HSA | **0.771 (0.991)** | **1.341 (1.754)** | 1.786 (2.204) | 2.122 (2.418) |
| | State | 0.968 (1.265) | 1.386 (1.780) | **1.746 (2.171)** | **2.046 (2.364)** |
| | | **MWIS** | | | |
| | HSA | **0.475 (0.729)** | **0.834 (1.326)** | **1.087 (1.654)** | 1.253 (1.786) |
| | State | 0.665 (1.081) | 0.899 (1.4380 | 1.104 (1.715) | 1.251 (1.832) |

**Table A.S4**. **Average forecast accuracy of HSA-level versus state-level models for three different influenza seasons (2022–2023, 2023–2024, and 2024–2025) using a naive baseline model.** Performance is summarized for one-, two-, three-, and four-week-ahead forecast horizons. Metrics include coverage rate, mean absolute percentage error (MAE), and mean weighted interval score (MWIS), each averaged across all locations and forecast weeks, within a season. Coverage rates closer to the nominal 95% level indicate better-calibrated uncertainty, while lower MAE and MWIS values indicate greater forecast accuracy. Bold values denote superior performance of HSA-level forecasts. Values in parentheses indicate standard deviations across locations and forecast weeks.

| Season | Level | Forecast horizon | | | |
|---|---|---|---|---|---|
| | | 1 | 2 | 3 | 4 |
| **2022/23** | | **Coverage Rate** | | | |
| | HSA | **0.871 (0.335)** | **0.800 (0.400)** | **0.752 (0.432)** | **0.724 (0.447)** |
| | State | 0.801 (0.400) | 0.769 (0.422) | 0.725 (0.447) | 0.691 (0.462) |
| | | **MAE** | | | |
| | HSA | **0.809 (1.103)** | **1.411 (1.849)** | 1.933 (2.415) | 2.385 (2.844) |
| | State | 1.048 (1.559) | 1.474 (1.938) | **1.904 (2.299)** | **2.309 (2.629)** |
| | | **MWIS** | | | |
| | HSA | **0.513 (0.810)** | **0.910 (1.381)** | **1.263 (1.826)** | 1.562 (2.152) |
| | State | 0.733 (1.332) | 0.994 (1.562) | 1.279 (1.826) | **1.553 (2.076)** |
| **2023/24** | | **Coverage Rate** | | | |
| | HSA | **0.955 (0.207)** | **0.923 (0.266)** | **0.908 (0.290)** | **0.900 (0.299)** |
| | State | 0.878 (0.328) | 0.882 (0.323) | 0.872 (0.334) | 0.866 (0.341) |
| | | **MAE** | | | |
| | HSA | **0.613 (0.805)** | **1.025 (1.321)** | **1.374 (1.662)** | **1.675 (1.867)** |
| | State | 0.882 (1.164) | 1.152 (1.487) | 1.425 (1.732) | 1.683 (1.868) |
| | | **MWIS** | | | |
| | HSA | **0.355 (0.547)** | **0.578 (0.913)** | **0.749 (1.166)** | **0.878 (1.292)** |
| | State | 0.558 (0.936) | 0.684 (1.140) | 0.814 (1.310) | 0.922 (1.391) |
| **2024/25** | | **Coverage Rate** | | | |
| | HSA | **0.850 (0.357)** | **0.789 (0.408)** | **0.738 (0.440)** | **0.695 (0.460)** |
| | State | 0.788 (0.409) | 0.746 (0.435) | 0.697 (0.460) | 0.656 (0.475) |
| | | **MAE** | | | |
| | HSA | **0.859 (1.080)** | **1.507 (1.797)** | 2.064 (2.237) | 2.529 (2.514) |
| | State | 1.052 (1.340) | 1.553 (1.836) | **2.036 (2.193)** | **2.472 (2.421)** |
| | | **MWIS** | | | |
| | HSA | **0.548 (0.803)** | **0.954 (1.370)** | **1.289 (1.707)** | **1.560 (1.950)** |
| | State | 0.726 (1.119) | 1.036 (1.494) | 1.336 (1.765) | 1.602 (1.937) |

**Table A.S5. Root mean square error (RMSE) between state- and HSA-level influenza emergency department (ED) trends for 173 Health Service Areas (HSAs).** HSAs are ordered from largest to smallest divergence (highest to lowest RMSE). We restricted analyses to the continental United States and to HSAs with complete ED visit percentage data observed between October 2022 and July 2025. To reduce stochastic noise, HSAs with populations below 250,000 were excluded; Washington, DC was also excluded because it is not a state and comprises a single HSA. The final analytic sample includes 31 states and 173 HSAs.

| State | HSA ID | HSA Description | RMSE |
|---|---|---|---|
| **Texas** | 405 | Potter (Amarillo), TX - Randall, TX | 3.492 |
| **Texas** | 538 | Webb (Laredo), TX - Zapata, TX | 3.194 |
| **Texas** | 427 | Hidalgo (Mcallen), TX - Starr, TX | 2.646 |
| **Texas** | 520 | Cameron (Brownsville), TX - Willacy, TX | 2.26 |
| **North Carolina** | 170 | Durham (Durham), NC - Granville, NC | 1.935 |
| **North Carolina** | 235 | Cabarrus (Concord), NC - Rowan, NC | 1.909 |
| **Texas** | 434 | Tarrant (Fort Worth), TX - Johnson, TX | 1.876 |
| **North Carolina** | 167 | Mecklenburg (Charlotte), NC - Union, NC | 1.813 |
| **North Carolina** | 225 | Buncombe (Asheville), NC - Henderson, NC | 1.712 |
| **Texas** | 505 | Brazoria (Lake Jackson), TX - Wharton, TX | 1.679 |
| **North Carolina** | 168 | New Hanover (Wilmington), NC - Brunswick, NC | 1.671 |
| **Minnesota** | 289 | St. Louis (Duluth), MN - Itasca, MN | 1.644 |
| **Tennessee** | 887 | Shelby (Memphis), TN - Tipton, TN | 1.583 |
| **Georgia** | 826 | Cherokee, GA - Douglas, GA | 1.522 |
| **Illinois** | 373 | Kane (Aurora), IL - DeKalb, IL | 1.502 |
| **South Carolina** | 244 | York (Rock Hill), SC - Chester, SC | 1.487 |
| **Georgia** | 154 | Floyd, GA - Bartow, GA | 1.454 |
| **Texas** | 413 | Jefferson (Beaumont), TX - Orange, TX | 1.378 |
| **Illinois** | 277 | Peoria (Peoria), IL - Tazewell, IL | 1.375 |
| **North Carolina** | 242 | Onslow, NC - Craven, NC | 1.322 |
| **Texas** | 415 | El Paso (El Paso), TX - Hudspeth, TX | 1.313 |
| **Texas** | 532 | Galveston, TX | 1.279 |
| **Minnesota** | 588 | Stearns (St. Cloud), MN - Benton, MN | 1.274 |
| **Indiana** | 902 | Vanderburgh (Evansville), IN - Warrick, IN | 1.255 |
| **Indiana** | 308 | Lake (Gary), IN - Porter, IN | 1.248 |
| **North Carolina** | 229 | Catawba (Hickory), NC - Burke, NC | 1.209 |
| **North Carolina** | 262 | Cumberland (Fayetteville), NC - Sampson, NC | 1.199 |
| **South Carolina** | 212 | Charleston (Charleston), SC - Berkeley, SC | 1.184 |
| **Texas** | 408 | Harris (Houston), TX - Fort Bend, TX | 1.183 |
| **Oklahoma** | 430 | Cleveland (Norman), OK - McClain, OK | 1.163 |
| **Virginia** | 5 | Newport News City, VA - Hampton City, VA | 1.151 |
| **Indiana** | 312 | St. Joseph (South Bend), IN - Marshall, IN | 1.141 |

| | | | |
|---|---|---|---|
| **Indiana** | 349 | Elkhart (Elkhart), IN - Kosciusko, IN | 1.133 |
| **Texas** | 437 | Nueces (Corpus Christi), TX - San Patricio, TX | 1.123 |
| **Indiana** | 300 | Tippecanoe (Lafayette), IN - Clinton, IN | 1.11 |
| **Tennessee** | 234 | Montgomery (Clarksville), TN - Stewart, TN | 1.11 |
| **North Carolina** | 218 | Gaston (Gastonia), NC - Cleveland, NC | 1.105 |
| **New York** | 86 | Orange (Newburgh), NY - Sullivan, NY | 1.099 |
| **Virginia** | 853 | Virginia Beach City, VA - Norfolk City, VA | 1.094 |
| **Tennessee** | 188 | Madison (Jackson), TN - Gibson, TN | 1.073 |
| **Texas** | 452 | Bell (Killeen), TX - Coryell, TX | 1.039 |
| **Indiana** | 390 | Hamilton, IN - Madison (Anderson), IN | 1.038 |
| **Michigan** | 296 | Muskegon (Muskegon), MI - Newaygo, MI | 1.032 |
| **Illinois** | 299 | Madison (Alton), IL - Jersey, IL | 1.022 |
| **New York** | 21 | Ontario (Geneva), NY - Wayne, NY | 1.022 |
| **Texas** | 406 | Lubbock (Lubbock), TX - Hockley, TX | 1.013 |
| **North Carolina** | 203 | Orange (Chapel Hill), NC - Harnett, NC | 1.011 |
| **Mississippi** | 146 | DeSoto, MS - Marshall, MS | 0.997 |
| **North Carolina** | 186 | Guilford (Greensboro), NC - Davidson, NC | 0.99 |
| **Texas** | 441 | Smith (Tyler), TX - Henderson, TX | 0.986 |
| **Illinois** | 279 | Champaign (Champaign), IL - Coles, IL | 0.985 |
| **Oregon** | 973 | Jackson (Medford), OR - Josephine, OR | 0.979 |
| **Georgia** | 829 | Clarke (Athens), GA - Barrow, GA | 0.972 |
| **New Jersey** | 64 | Atlantic (Atlantic City), NJ - Cape May, NJ | 0.966 |
| **Michigan** | 285 | Ingham (Lansing), MI - Eaton, MI | 0.965 |
| **North Carolina** | 149 | Forsyth (Winston-Salem), NC - Surry, NC | 0.941 |
| **Kentucky** | 2 | Kenton (Covington), KY - Campbell, KY | 0.934 |
| **North Carolina** | 195 | Pitt, NC - Beaufort, NC | 0.928 |
| **Texas** | 462 | McLennan (Waco), TX - Hill, TX | 0.923 |
| **West Virginia** | 7 | Kanawha (Charleston), WV - Putnam, WV | 0.918 |
| **Wisconsin** | 278 | Brown (Green Bay), WI - Oconto, WI | 0.907 |
| **Colorado** | 711 | Mesa, CO - Garfield, CO | 0.901 |
| **Georgia** | 830 | Richmond (South Augusta), GA - Columbia, GA | 0.898 |
| **Minnesota** | 540 | Hennepin (Minneapolis), MN - Anoka, MN | 0.895 |
| **Colorado** | 760 | Weld, CO - Morgan, CO | 0.879 |
| **Wisconsin** | 355 | Sheboygan (Sheboygan), WI - Manitowoc, WI | 0.879 |
| **Illinois** | 325 | St. Clair (Belleville), IL - Clinton, IL | 0.877 |
| **New York** | 41 | Westchester (Yonkers), NY - Dutchess, NY | 0.87 |
| **Wisconsin** | 382 | Racine (Racine), WI - Kenosha, WI | 0.867 |
| **Texas** | 510 | Hays (San Marcos), TX - Caldwell, TX | 0.864 |

| | | | |
|---|---|---|---|
| **Kansas** | 624 | Johnson (Overland Park), KS - Douglas, KS | 0.862 |
| **Kentucky** | 272 | Jefferson (Louisville), KY - Bullitt, KY | 0.86 |
| **Tennessee** | 147 | Knox (Knoxville), TN - Blount, TN | 0.848 |
| **Texas** | 464 | Brazos (Bryan), TX - Washington, TX | 0.847 |
| **New Mexico** | 732 | Dona Ana (Las Cruces), NM - Luna, NM | 0.846 |
| **Maryland** | 48 | Frederick, MD | 0.839 |
| **Illinois** | 303 | Will (Joliet), IL - Grundy, IL | 0.836 |
| **Illinois** | 318 | Sangamon (Springfield), IL - Christian, IL | 0.831 |
| **Georgia** | 157 | Hall, GA - Habersham, GA | 0.829 |
| **Michigan** | 322 | Saginaw (Saginaw), MI - Tuscola, MI | 0.828 |
| **New York** | 865 | Rockland, NY | 0.826 |
| **Virginia** | 14 | Roanoke City, VA - Roanoke, VA | 0.82 |
| **South Carolina** | 246 | Horry, SC - Georgetown, SC | 0.817 |
| **Michigan** | 375 | Jackson (Jackson), MI - Lenawee, MI | 0.808 |
| **Massachusetts** | 32 | Hampden (Springfield), MA - Hampshire, MA | 0.804 |
| **North Carolina** | 198 | Wake (Raleigh), NC - Johnston, NC | 0.804 |
| **New York** | 54 | Erie (Buffalo), NY - Monroe, NY | 0.792 |
| **Oregon** | 782 | Lane (Eugene), OR - Douglas, OR | 0.786 |
| **Georgia** | 894 | Coweta, GA - Troup (La Grange), GA | 0.783 |
| **Michigan** | 320 | Ottawa (Holland), MI - Allegan, MI | 0.778 |
| **Michigan** | 327 | Berrien (Benton Harbor), MI - Van Buren, MI | 0.776 |
| **Texas** | 425 | Travis (Austin), TX - Williamson, TX | 0.773 |
| **Indiana** | 304 | Allen (Fort Wayne), IN - Noble, IN | 0.772 |
| **Wisconsin** | 306 | Winnebago (Oshkosh), WI - Fond du Lac, WI | 0.765 |
| **New York** | 80 | St. Lawrence, NY - Jefferson, NY | 0.756 |
| **Tennessee** | 231 | Sumner (Hendersonville), TN - Wilson, TN | 0.754 |
| **Michigan** | 831 | Washtenaw, MI - Livingston, MI | 0.751 |
| **Georgia** | 893 | Muscogee (Columbus), GA - Harris, GA | 0.75 |
| **South Carolina** | 184 | Florence (Florence), SC - Darlington, SC | 0.739 |
| **South Carolina** | 182 | Greenville (Greenville), SC - Anderson, SC | 0.732 |
| **Mississippi** | 456 | Harrison (Biloxi), MS - Hancock, MS | 0.723 |
| **Iowa** | 545 | Linn (Cedar Rapids), IA - Johnson, IA | 0.722 |
| **New Jersey** | 23 | Camden (Camden), NJ - Burlington, NJ | 0.711 |
| **Maine** | 9 | Cumberland (Portland), ME - Knox, ME | 0.703 |
| **Texas** | 410 | Bexar (San Antonio), TX - Guadalupe, TX | 0.703 |
| **Virginia** | 109 | Prince William (Dale City), VA - Fauquier, VA | 0.701 |
| **Wisconsin** | 282 | Marathon (Wausau), WI - Wood, WI | 0.691 |
| **Mississippi** | 411 | Hinds (Jackson), MS - Rankin, MS | 0.69 |

| | | | |
|---|---|---|---|
| **Kansas** | 576 | Sedgwick (Wichita), KS - Butler, KS | 0.683 |
| **New York** | 88 | Saratoga, NY - Schenectady (Schenectady), NY | 0.678 |
| **New Jersey** | 126 | Mercer, NJ | 0.67 |
| **Nevada** | 962 | Washoe (Reno), NV - Carson City, NV | 0.663 |
| **Minnesota** | 941 | Olmsted (Rochester), MN - Winona, MN | 0.651 |
| **Oregon** | 719 | Deschutes, OR - Crook, OR | 0.64 |
| **Tennessee** | 211 | Rutherford (Murfreesboro), TN - Warren, TN | 0.634 |
| **Texas** | 495 | Denton (Denton), TX - Wise, TX | 0.633 |
| **Kansas** | 554 | Shawnee (Topeka), KS - Riley, KS | 0.627 |
| **Virginia** | 69 | Fairfax, VA - Arlington, VA | 0.615 |
| **Virginia** | 99 | Albemarle, VA - Charlottesville City, VA | 0.608 |
| **Illinois** | 291 | Winnebago (Rockford), IL - Ogle, IL | 0.606 |
| **New York** | 10 | Albany (Albany), NY - Rensselaer, NY | 0.602 |
| **Idaho** | 734 | Kootenai, ID - Bonner, ID | 0.592 |
| **Michigan** | 328 | Genesee (Flint), MI - Lapeer, MI | 0.588 |
| **Maryland** | 875 | Harford, MD - Cecil, MD | 0.586 |
| **New Jersey** | 87 | Morris (Parsippany-Troy Hills Township), NJ - Sussex, NJ | 0.586 |
| **Nebraska** | 561 | Lancaster (Lincoln), NE - Gage, NE | 0.574 |
| **Maine** | 17 | Penobscot (Bangor), ME - Hancock, ME | 0.57 |
| **Virginia** | 135 | Stafford, VA - Spotsylvania, VA | 0.553 |
| **New York** | 56 | Onondaga (Syracuse), NY - Oswego, NY | 0.552 |
| **New York** | 59 | Oneida (Utica), NY - Madison, NY | 0.543 |
| **Texas** | 453 | Dallas (Dallas), TX - Collin, TX | 0.539 |
| **Wisconsin** | 280 | Milwaukee (Milwaukee), WI - Waukesha, WI | 0.539 |
| **Michigan** | 309 | Kent (Grand Rapids), MI - Ionia, MI | 0.516 |
| **Tennessee** | 148 | Davidson (Nashville-Davidson), TN - Williamson, TN | 0.514 |
| **Virginia** | 33 | Henrico (Richmond), VA - Chesterfield, VA | 0.506 |
| **Tennessee** | 885 | Hamilton (Chattanooga), TN - Marion, TN | 0.505 |
| **Michigan** | 348 | Kalamazoo (Kalamazoo), MI - St. Joseph, MI | 0.498 |
| **Massachusetts** | 68 | Bristol (New Bedford), MA | 0.488 |
| **New Mexico** | 693 | Bernalillo (Albuquerque), NM - Sandoval, NM | 0.488 |
| **Indiana** | 275 | Marion (Indianapolis), IN - Johnson, IN | 0.487 |
| **Iowa** | 546 | Polk (Des Moines), IA - Dallas, IA | 0.472 |
| **Wisconsin** | 301 | Dane (Madison), WI - Sauk, WI | 0.467 |
| **Michigan** | 274 | Wayne (Detroit), MI - Oakland, MI | 0.462 |
| **South Carolina** | 160 | Richland (Columbia), SC - Lexington, SC | 0.46 |
| **Illinois** | 287 | Cook (Chicago), IL - DuPage, IL | 0.443 |

| | | | |
|---|---|---|---|
| **Delaware** | 3 | Sussex, DE - Kent, DE | 0.44 |
| **Utah** | 744 | Davis, UT - Weber (Ogden), UT | 0.435 |
| **Vermont** | 49 | Chittenden (Burlington), VT - Franklin, VT | 0.435 |
| **Nebraska** | 542 | Douglas (Omaha), NE - Sarpy, NE | 0.42 |
| **Utah** | 703 | Utah (Provo), UT - Sanpete, UT | 0.394 |
| **Georgia** | 825 | Henry, GA - Forsyth, GA | 0.389 |
| **Massachusetts** | 101 | Worcester (Worcester), MA - Franklin, MA | 0.383 |
| **Georgia** | 190 | Cobb (Marietta), GA | 0.381 |
| **Georgia** | 153 | Fulton (Atlanta), GA - DeKalb, GA | 0.376 |
| **New Jersey** | 66 | Essex (Newark), NJ - Middlesex, NJ | 0.371 |
| **Kentucky** | 18 | Fayette (Lexington-Fayette), KY - Jessamine, KY | 0.367 |
| **Oklahoma** | 417 | Oklahoma (Oklahoma City), OK - Canadian, OK | 0.345 |
| **Colorado** | 796 | Larimer, CO | 0.341 |
| **New York** | 83 | Suffolk, NY - Nassau, NY | 0.339 |
| **Colorado** | 754 | El Paso (Colorado Springs), CO - Teller, CO | 0.337 |
| **Colorado** | 795 | Boulder, CO - Broomfield, CO | 0.334 |
| **Idaho** | 716 | Ada (Boise City), ID - Canyon, ID | 0.327 |
| **Massachusetts** | 74 | Essex (Lynn), MA | 0.32 |
| **Delaware** | 75 | New Castle (Wilmington), DE | 0.317 |
| **New Jersey** | 108 | Monmouth, NJ - Ocean (Brick Township), NJ | 0.309 |
| **Massachusetts** | 22 | Middlesex, MA - Suffolk (Boston), MA | 0.295 |
| **Oregon** | 689 | Multnomah (Portland), OR - Washington, OR | 0.277 |
| **New Jersey** | 36 | Bergen, NJ - Hudson (Jersey City), NJ | 0.236 |
| **Nevada** | 707 | Clark (Las Vegas), NV - Nye, NV | 0.217 |
| **Maryland** | 869 | Montgomery, MD - Prince Georges, MD | 0.208 |
| **Maryland** | 16 | Baltimore, MD - Baltimore City, MD | 0.191 |
| **Colorado** | 688 | Denver (Denver), CO - Jefferson, CO | 0.159 |
| **Utah** | 708 | Salt Lake (Salt Lake City), UT - Tooele, UT | 0.141 |

### Supplementary Material B — HSA Size Threshold

For each location $i$ (HSA or state) and week $t = 1, \ldots, T$, let $y_{i,t}$ denote the weekly percentage of ED visits attributable to influenza.

#### Overall relative variability

We quantified week-to-week variability using the coefficient of variation (CV):

$$\mathrm{CV}_i = \frac{\mathrm{sd}(y_{i,1:T})}{\mathrm{mean}(y_{i,1:T})}.$$

This normalizes variability by the typical level of influenza activity, enabling comparisons across locations with different baseline incidence. Larger values of the CV indicate greater relative fluctuation around the mean, reflecting greater week-to-week variability in influenza ED visits percentages.

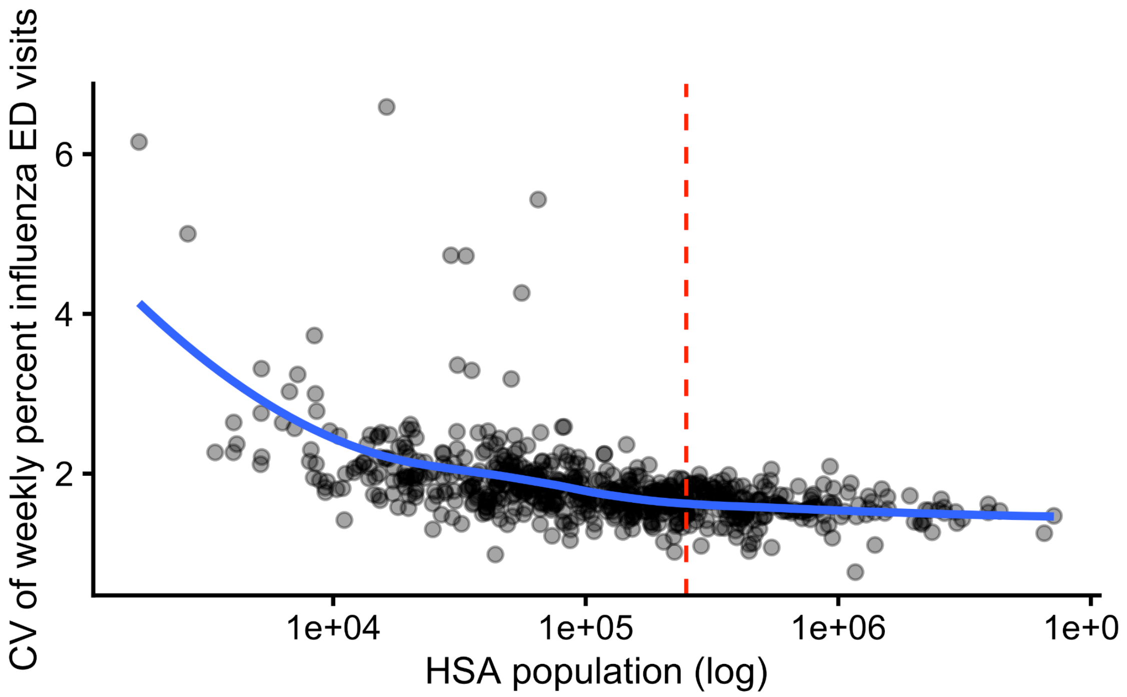


**Fig. B.S1. Relationship between Health Service Area (HSA) population size and week-to-week variability in the percentage of emergency department (ED) visits attributable to influenza.** Variability is measured using the coefficient of variation (CV). The smooth curve shows declining relative variability with increasing population size, followed by a plateau near 250,000 residents, indicating diminishing reductions in stochastic noise beyond this threshold. This pattern motivates the cutoff used in the main analysis.

#### Residual week-to-week variability

Because overall CV reflects both epidemic dynamics (e.g., seasonal peaks) and stochastic fluctuations, we also quantified short-term variability after removing smooth temporal trends.
For each location $i$, we fit a generalized additive model (GAM) with a spline over time :

$$y_{i,t} = \beta_{0,i} + f_i(t) + \varepsilon_{i,t},$$

where $f_i(t)$ is a smooth function of the week (fit using REML). Let $\hat{\mu}_{i.t}$ denote the fitted mean trajectory.

Residuals were defined as $r_{i,t} = y_{i,t} - \hat{\mu}_{i,t}$, and residual relative variability was calculated as

$$\mathrm{CV}_i^{\mathrm{resid}} = \frac{\mathrm{sd}(r_i, 1:T)}{\mathrm{mean}(\hat{\mu}_{i,1:T})}.$$

This measure captures short-term fluctuations in weekly influenza ED visit percentages after removing smooth seasonal patterns. Larger values of the residual CV indicate greater irregular week-to-week variability beyond the underlying epidemic trend, whereas smaller values reflect more stable deviations around the fitted trajectory.

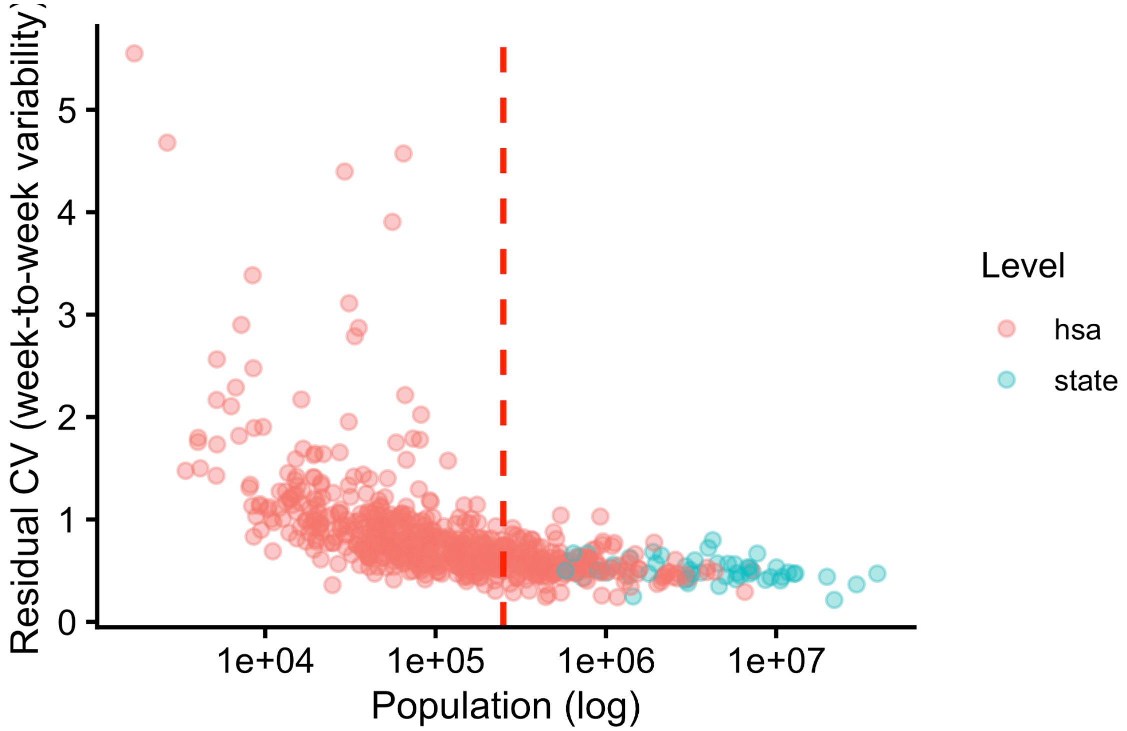


**Fig. B.S2. Relationship between population size and residual week-to-week variability in influenza ED visit percentages after removing smooth temporal trends.** Variability declines sharply with population size and plateaus near 250,000 residents, supporting the population threshold used in the main analysis.

### Supplementary Material C — Covariate Selection and Alternative Inference Models

This supplement provides covariate selection procedures, and additional model fitting results and diagnostic analyses that support the findings presented in the main text. In the main analysis, we focus on modeling log-transformed differences in weighted interval scores as a function of the log HSA–state population ratio, proportion of urban population and the number of MSAs per state. Here, we extend this analysis by presenting diagnostic plots, robustness checks across alternative modeling approaches, and additional model specifications that were not included in the main script.

#### Covariate Selection and Model Specification

To characterize associations between regional covariates and the average difference in WIS between HSA- and state-level forecasts, we focused on variables representing population size and urbanization, motivated by prior work highlighting their roles in influenza transmission dynamics and spatial heterogeneity (1, 2) .

We represented population size using both the HSA–state population ratio and separate measures of HSA and state population size, allowing us to assess how different parameterizations capture aggregation effects. For urbanization, we evaluated multiple representations, including the proportion of residents living in urban-designated areas (urban share), population density, and the proportion of land area classified as urban, each capturing distinct dimensions of urban structure.

We additionally considered covariates reflecting demographic composition and spatial structure. Age composition variables were included as proxies for differences in transmission potential, given well-established heterogeneity in contact patterns across age groups, particularly among children (3–5). State-level structural variables, such as total land area and the number of large metropolitan regions, were included to capture heterogeneity in epidemic dynamics that may be more pronounced in geographically large and heterogeneous states (6). In particular, we included the number of Metropolitan Statistical Areas (MSAs) to represent the distribution of major population centers. We hypothesized that MSA count would better capture spatial heterogeneity in transmission than HSA count, as MSAs reflect urban population centers and commuting networks, whereas HSAs are defined primarily by healthcare referral patterns.

Covariate selection followed a targeted, theory-driven sensitivity analysis. We began with a set of candidate variables motivated by prior literature, including multiple representations of population size, urbanization, demographic composition, and spatial structure. We evaluated candidate variables by fitting models across forecast horizons while varying covariate combinations and functional forms. As part of this process, we performed stepwise screening to identify variables with statistically meaningful contributions at shorter horizons. Final variable selection was guided by consistency of effect direction, interpretability, and stability across model specifications and forecast horizons. We additionally included an interaction between population ratio and urban share to capture potential joint effects of population concentration and urbanization on forecast performance, as these variables are conceptually related. At the one-week horizon, we prioritized variables that were statistically significant with coherent effect directions. At longer horizons, we focused less on significance and more on directional consistency given increased uncertainty. To avoid redundancy, highly correlated predictors were not included simultaneously, resulting in a parsimonious final specification.

The final model includes the HSA–state population ratio, urban share, their interaction, and MSA count. MSA count was selected as an interpretable proxy for spatial fragmentation and showed robust associations across specifications.

Population and demographic variables were derived from 2020 U.S. Census data, while metropolitan delineations were obtained from the Office of Management and Budget (OMB). Census data were accessed using the *tidycensus* R package (7, 8), with code available in the accompanying GitHub repository (9).

To improve model stability and interpretability, we applied log transformations to skewed covariates (e.g., population ratio and urban share). The response variable (difference in WIS) was also log-transformed after applying a constant shift to accommodate negative values.

We evaluated three modeling approaches: generalized linear models (GLM), generalized additive models (GAM), and Bayesian additive regression trees (BART). All approaches yielded qualitatively similar patterns. BART showed modest gains in predictive accuracy at shorter forecast horizons, but this advantage diminished at longer horizons, where GLM and GAM produced more stable and interpretable results. For consistency, all models were fit using the same covariates. To balance interpretability and predictive stability, we focus on GLM results and use GAM and BART as robustness checks; full results and diagnostics are provided in Supplementary Material C.

The GLM models the expected average difference in WIS between HSA- and state-level forecasts as a function of the selected covariates. Let $y_i^h$ denote the average difference in WIS for HSA $i$ at forecast horizon $h$, aggregated across seasons and forecast dates as defined in Equation (7), $\Delta \mathrm{MWIS}_i^h$, and let $s(i)$ denote the state containing HSA $i$. The HSA–state ratio is the ratio of HSA to state population, urban share is the proportion of the HSA population residing in urban-designated areas, and MSA count denotes the number of Metropolitan Statistical Areas in the state. Because $y_i^h$ can take negative values, we apply a constant shift ( $c = 0.01$) prior to log transformation. The final model is a Gaussian linear regression on the transformed outcome:

$$\log\Big(y_i^h - \min_i y_i^h + c\Big) = \beta_0 + \beta_1 \log(R_i) + \beta_2 \log(U_i) + \beta_3 \log(R_i)\log(U_i) + \beta_4 M_{s(i)} + \varepsilon_i,$$

$$\varepsilon_i \sim N(0, \sigma^2).$$

where $R_i$ denotes the HSA-state population ratio, $U_i$ denotes the proportion of the HSA population residing in urban-designated areas, and $M_{s(i)}$ denotes the number of MSAs in the corresponding state. Models were fit separately for each forecast horizon (1-3 weeks ahead); for clarity, the horizon index is omitted. Horizon-specific coefficient estimates are reported in Supplementary Table C.S1.

Finally, we note that the assumption of independent, homoscedastic errors is unlikely to hold exactly given shared data sources, spatial proximity, and common modeling structure across

HSAs. Accordingly, results should be interpreted as descriptive associations rather than causal effects.

#### Diagnostics & model comparison

Section C.1 presents diagnostic plots and effect summaries for the model specification used in the main text, alongside comparisons with alternative modeling approaches, including generalized additive models (GAMs) and Bayesian additive regression trees (BART). These diagnostics assess model fit, distributional assumptions, and the consistency of fitted relationships across modeling frameworks.

**(A)**

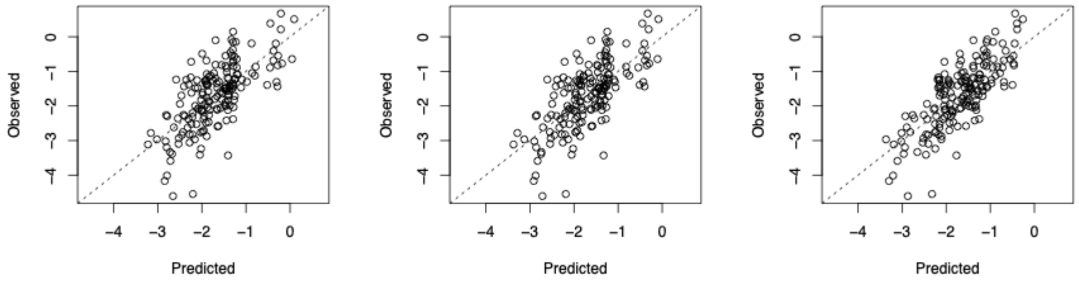


**(B)**

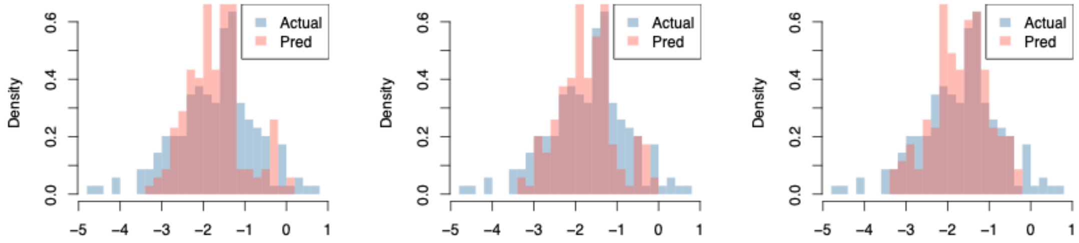


**(C)**

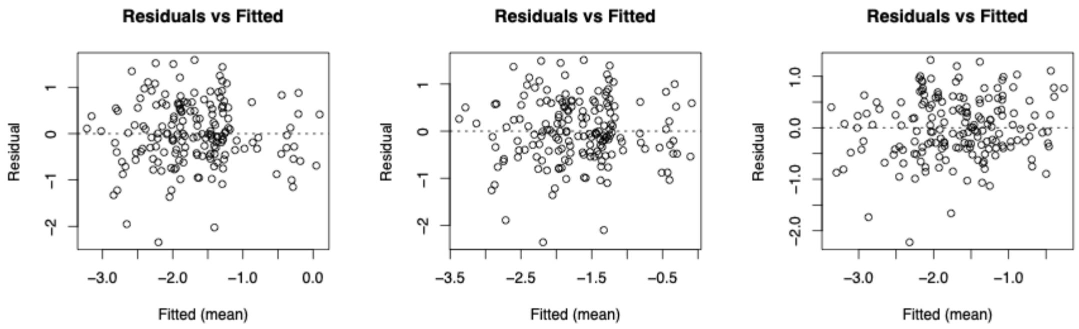

(D)

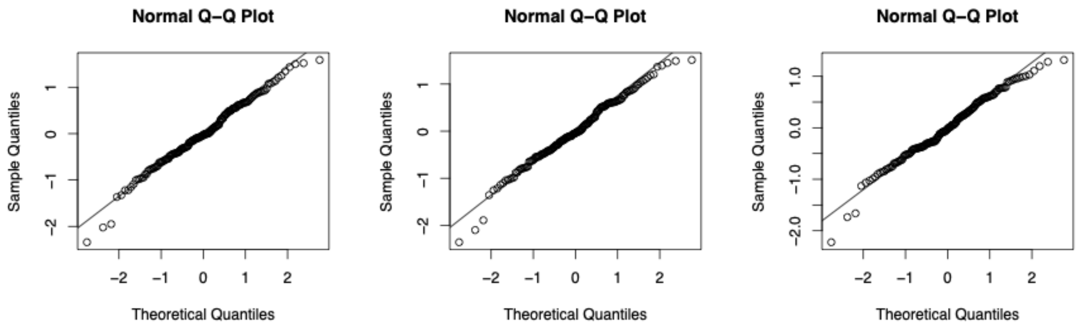
Normal Q–Q Plot
Normal Q–Q Plot
Normal Q–Q Plot
Sample Quantiles
Theoretical Quantiles

(E)

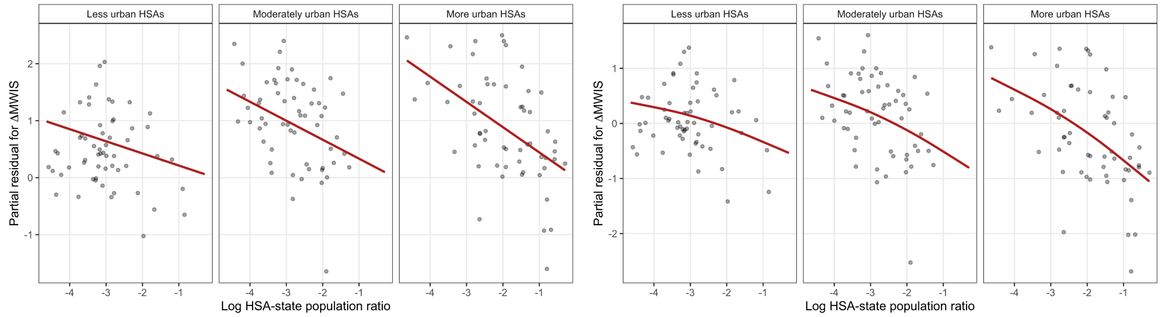
Less urban HSAs
Moderately urban HSAs
More urban HSAs
Less urban HSAs
Moderately urban HSAs
More urban HSAs
Partial residual for ΔMWIS
Log HSA-state population ratio
Log HSA-state population ratio

(F)

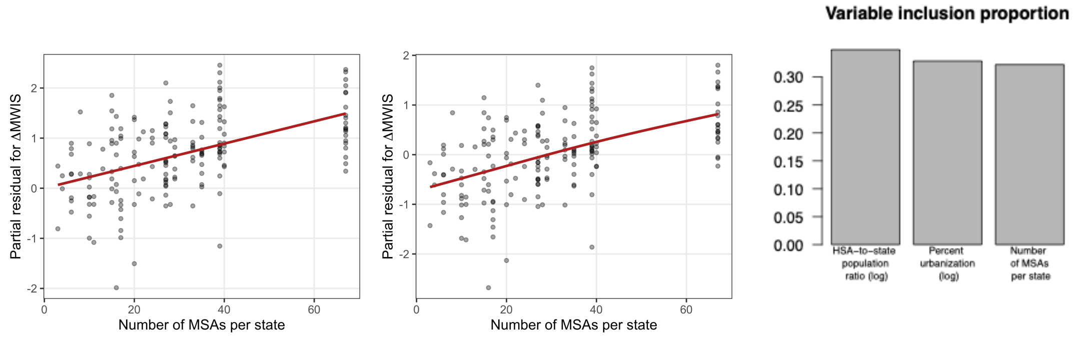
Partial residual for ΔMWIS
Number of MSAs per state
Partial residual for ΔMWIS
Number of MSAs per state
Variable inclusion proportion
HSA–to–state population ratio (log)
Percent urbanization (log)
Number of MSAs per state

**Fig. C.S1. Model diagnostics and robustness checks across modeling approaches.** Diagnostic plots and effect summaries comparing three modeling approaches used to relate population structure to differences in forecasting performance. Columns correspond to generalized linear models (GLM; left), generalized additive models (GAM; middle), and Bayesian additive regression trees (BART; right). Across all three approaches, models relate the log-transformed difference in weighted interval score (log(shifted ΔWIS)) to the log HSA–state population ratio and the number of MSAs per state. (A) Observed versus predicted values of log(shifted ΔWIS. (B) Density comparisons of observed and model-predicted log(shifted ΔWIS). (C) Residuals versus fitted values for log(shifted ΔWIS. (D) Normal Q–Q plots for log(shifted ΔWIS), assessing model fit and distributional assumptions. (E) Partial residual plots stratified by terciles of HSA urbanization. Red lines show the predicted relationship at the median urbanization level within each tercile, holding other covariates constant. The negative association between HSA-state population and forecast improvement is strongest in more urbanized HSAs. (F) Marginal effects of key predictors on log(shifted ΔWIS). Across models, fitted relationships and diagnostics are broadly consistent, indicating that the main conclusions are robust to model specification; for BART, variable inclusion proportions are shown instead of parametric effect estimates.

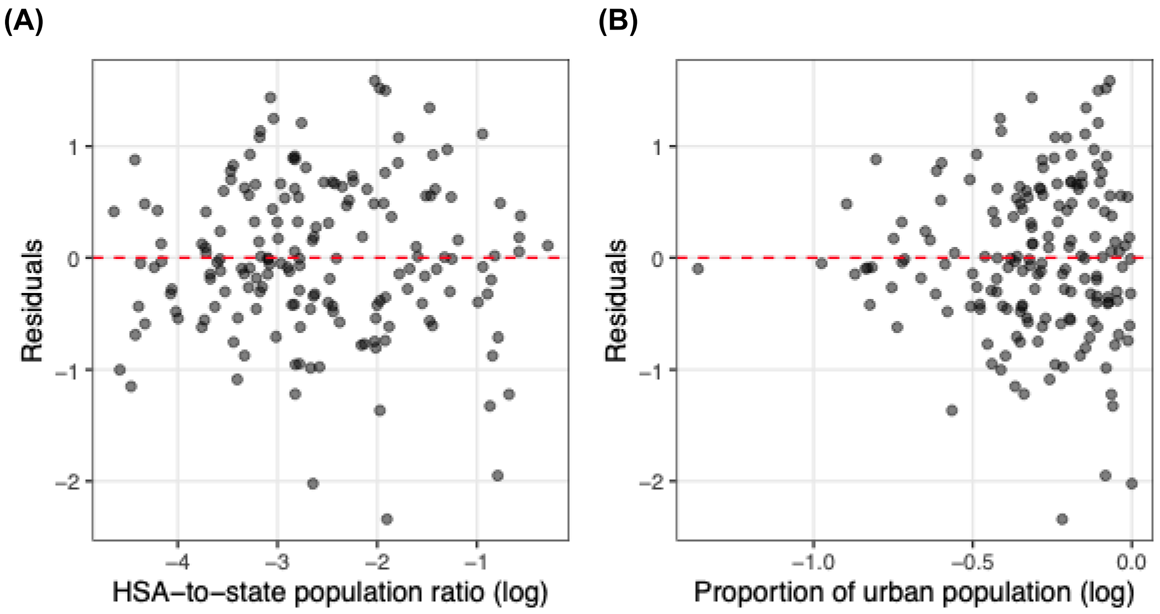


**Fig. C.S2. Diagnostic plots of residuals versus predictor variables for GLM .** Residuals are plotted against the primary predictors to assess the appropriateness of the model's linear specification. In both panels, the dashed red line represents the zero-residual baseline. (A) HSA−to−state population ratio (log): The random scatter of points around the zero line confirms that the linear functional form for the population ratio is appropriate and exhibits no systematic bias or non-linearity (B) Proportion of urban population (log): Although variance increases at higher levels of urbanization (heteroscedasticity), the residuals remain centered around the zero line, indicating that the model’s central tendency is correctly captured across the full range of urbanization values.

**Table C.S1. Performance and coefficients of the generalized linear model (GLM) relating population structure to local forecasting gains.** Estimated coefficients from the generalized linear model relating differences in weighted interval score (ΔWIS) to HSA-level and state-level population characteristics, fitted separately for each forecasting horizon using the full dataset.

| Forecast Horizon (weeks) | Predictor Variables | | | | | Adjusted $R^2$ |
|---|---|---|---|---|---|---|
| | **Intercept** | **HSA-state population ratio (log)** | **Proportion of urban population (log)** | **Number of MSAs state** | **HSA-state population ratio(log): proportion of urban population (log)** | |
| **1** | -3.568*** | -0.489*** | -1.421* | 0.022*** | -0.560* | 0.4676 |
| **2** | -2.838*** | -0.272** | -0.417 | 0.019*** | -0.344 • | 0.3327 |
| **3** | -2.152*** | -0.138 • | -0.073 | 0.013*** | -0.167 | 0.1874 |
| • p<0.1, * p<0.05, ** p<0.01, *** p<0.001 | | | | | | |

**Table C.S2.** Five-fold cross-validated predictive performance of generalized linear models (GLM), generalized additive models (GAM), and Bayesian additive regression trees (BART) for forecasting horizons of 1–3 weeks. Performance is evaluated using the coefficient of determination ($R^2$), root mean squared error (RMSE), and mean absolute error (MAE), all computed on the log-transformed, shifted difference in weighted interval score (log(shifted ΔWIS)). Results indicate comparable predictive accuracy across modeling approaches, with modest declines in performance at longer forecasting horizons.

Performance is assessed using cross-validated $R^2$, RMSE, and MAE computed via five-fold cross-validation.

| Forecast horizon (weeks) | Model | $R^2$ (CV) | RMSE (CV) | MAE (CV) |
|---|---|---|---|---|
| **1** | GLM | 0.4441 | **0.7062** | **0.5529** |
| | GAM | 0.4396 | **0.7090** | **0.5593** |
| | BART | **0.4530** | **0.7005** | 0.5655 |
| **2** | GLM | **0.3006** | 0.6614 | **0.5065** |
| | GAM | **0.3039** | **0.6598** | **0.5053** |

| | | | | |
|---|---|---|---|---|
| | BART | 0.2832 | 0.6696 | **0.5090** |
| **3** | GLM | **0.1609** | **0.5801** | **0.4204** |
| | GAM | 0.1560 | **0.5818** | **0.4240** |
| | BART | 0.1477 | **0.5846** | **0.4279** |

### Supplementary Material D — Forecasting Models

#### Gradient Boosting Quantile Regression (GBQR)

All primary forecasts were generated using a Gradient Boosting Quantile Regression (GBQR) framework (10), which models conditional quantiles of the target variable (e.g., percent of $y_{i,t+h}$ influenza-related ED visits) given features $x_{i,t}$.

Formally, for each forecast horizon $h$ and quantile level $\tau$, the model estimates

$$\hat{q}_{\tau}^{(h)}(x_{i,t}) = \sum_{m=1}^{M} f_{m,\tau}^{(h)}(x_{i,t})$$

where $\hat{q}_{\tau}^{(h)}(x_{i,t})$ denotes the predicted $\tau$-th conditional quantile of the future outcome $y_{i,t+h}$ and $f_{m,\tau}^{(h)}(\cdot)$ denotes the regression trees added sequentially by gradient boosting.

Model parameters are optimized to minimize the pinball (quantile) loss:

$$\mathcal{L}_{\tau}^{(h)} = \sum_{t} \rho_{\tau}\Big(y_{i,t+h} - f_{\tau}^{(h)}(x_{i,t})\Big),$$

$$\rho_{\tau} = u(\tau - \mathbf{1}\{u < 0\}),$$

which encourages accurate estimation of the conditional $\tau$-th quantile of the predictive distribution.

Each quantile level is estimated separately using a single model fit to pooled data across all locations, and forecasts are ensembled across 100 random training subsamples (bagging) to improve stability.

The final predictive distribution is defined by the set of quantile forecasts $\{\hat{q}_{\tau}^{(h)}\}$; we summarize results using the median (50th percentile) and 95% prediction intervals (2.5th and 97.5th percentiles).

Models were trained using the LightGBM implementation of quantile regression (11) in Python with default hyperparameters (e.g., num_leaves = 31, learning_rate = 0.1, n_estimators = 100), with the quantile level (alpha) specified for each model.

#### Autoregressive Integrated Moving Average (ARIMA) Model

We implemented an automated autoregressive integrated moving average (ARIMA) model as a benchmark forecasting approach. For each location $l$, we fit an ARIMA model to weekly percent influenza ED visits using the experimental design described in the main Methods section (Forecasting Experimental Design). Specifically, models were trained using all observations prior to the forecast date within the target season, together with all observations from the other two seasons.

Model orders were selected automatically using the auto.arima() function from the **forecast** R package (12), which determines the autoregressive ($p$), differencing ($d$), and moving-average ( $q$) orders by minimizing the corrected Akaike Information criterion (AICc).

Let $y_t$ denote the observed weekly outcome at time $t$. The ARIMA model is defined as:

$$\Phi_p(B)(1-B)^d y_t = \Theta_q(B)\varepsilon_t,$$

where, $B$ is the backshift operator, $\phi(B)$ and $\theta(B)$ are polynomials of order $p$ and $q$, respectively, and $\epsilon_t \sim \mathcal{N}(0, \sigma^2)$ are Gaussian innovations.

Forecasts up to four weeks ahead were generated for each location $l$. To obtain probabilistic forecasts, we simulated 2000 future trajectories from the fitted ARIMA model using stats::simulate(), and empirical quantiles were computed at each forecast horizon. Negative simulated values were truncated at zero. All model fitting and forecasting procedures are available in the GitHub repository (13).

#### Naive Quantile Baseline (FluSight baseline)

We implemented a naive quantile baseline model as an additional benchmark. This baseline corresponds to the quantile baseline used in the U.S. CDC FluSight forecasting framework (14) and implemented in the Flu-Metrocast framework (14, 15).

For each location $l$, the baseline model was fit to weekly percent influenza ED visits using the same retrospective training framework described in the main text.

Let $y_t$ denote the observed weekly outcome at time $t$, and define weekly changes as

$$\Delta y_t = y_t - y_{t-1}.$$

The baseline constructs a predictive distribution from the empirical distribution of historical weekly changes ($\Delta y_t$), without explicitly modeling temporal autocorrelation or seasonal structure. Future trajectories are generated by sampling independently from this empirical distribution and accumulating sampled changes over the forecast horizon.

To obtain probabilistic forecasts, we generated 1000 Monte Carlo samples from the baseline predictive distribution and computed empirical quantiles at each forecast horizon. All model fitting and forecasting procedures are available in the GitHub repository (13).

### Supplementary Material E — Performance Metrics

#### Evaluation Metrics

To compare the performance of HSA-level and state-level forecasts when evaluated against the HSA level observations, we use three standard metrics: 95% Prediction Interval Coverage Rate (Coverage Rate), Mean Absolute Error (MAE), and Mean Weighted Interval Score (MWIS). Together, these capture both point forecast accuracy and probabilistic calibration.

Coverage measures the proportion of observed values that fall within the forecasted 95% prediction interval (2.5th-97.5th percentiles). For each forecasted week, we assign a value of 1 if the observed value falls within the interval and 0 otherwise:

$$\text{Coverage Rate}_i^h = \frac{1}{T}\sum_{t=1}^{T} 1\big(l_{i,t}^h \le y_{i,t+h} \le u_{i,t}^h\big) \quad . \tag{2}$$

Where $y_{i,t}$ is the observed value and $l_{i,t}^h, u_{i,t}^h$ are the lower and upper interval bounds at location $i$, time $t$, forecast horizon $h$, and $\mathbf{1}(\cdot)$ is an indication function. Here,

$i$ indexes a geographic unit (either an HSA or a state), with $s(i)$ denoting the state corresponding to HSA $i$. Values closer to the specified confidence level (i.e., 0.95) indicate better calibration of forecast uncertainty.

MAE quantifies the average absolute difference between the predicted median ($\hat{y}_{i,t+h}$) and the observed value ($y_{i,t+h}$), here $h$ denotes the forecast horizon:

$$\text{MAE}_i^h = \frac{1}{T}\sum_{t=1}^{T} |y_{i,t+h} - \hat{y}_{i,t+h}| \quad . \tag{3}$$

Here, $i$ indexes a geographic unit (either an HSA or a state), with $s(i)$ denoting the state corresponding to HSA $i$. Lower MAE values indicate more accurate point forecasts.

We assess probabilistic forecast performance using the mean weighted interval score (MWIS). The weighted interval score (WIS) (16) is a proper scoring rule defined for a single forecast-observation pair. WIS evaluates the full predictive distribution by combining information on accuracy, sharpness, and calibration across multiple quantile levels:

$$\text{WIS}_{\alpha\{0:K\}}(F, y) = \frac{1}{2K+1} \times \sum_{k=1}^{2K+1} 2 \times \{1(y \le q_{\tau_k}) - \tau_k\} \times (q_{\tau_k} - y) \quad . \tag{4}$$

Where $F$ denotes the forecast distribution and $q_{\tau_k} = F^{-1}(\tau_k)$ is the predicted quantile at level $\tau_k$. We use the standard set of quantile levels $\tau_k \in \{0.01, 0.025, 0.05, \cdots, 0.95, 0.975, 0.99\}$ following the CDC FluSight evaluation framework. WIS is closely related to the pinball loss and can be expressed as a weighted sum of pinball losses across these quantiles.

For each HSA $i$ and forecast horizon $h$, we compute the mean weighted interval score (MWIS) by averaging WIS values across all forecast dates, where lower MWIS values indicate better overall probabilistic forecast performance. Formally, MWIS is defined as:

$$\mathrm{MWIS}_i^h = \frac{1}{T}\sum_{t=1}^{T} \mathrm{WIS}_{i,t}^h \quad . \tag{5}$$

Here, $i$ indexes a geographic unit (either an HSA or a state), and $s(i)$ denotes the state corresponding to HSA $i$, as defined above.

8. US Census Bureau, Decennial Census of Population and Housing.
